# Cell autonomous microglia defects in a stem cell model of frontotemporal dementia

**DOI:** 10.1101/2024.05.15.24307444

**Authors:** Abhirami K. Iyer, Lisa Vermunt, Farzaneh S. Mirfakhar, Miguel Minaya, Mariana Acquarone, Rama Krishna Koppisetti, Arun Renganathan, Shih-Feng You, Emma P. Danhash, Anthony Verbeck, Grant Galasso, Scott M. Lee, Jacob Marsh, Alissa L. Nana, Salvatore Spina, William W. Seeley, Lea T. Grinberg, Sally Temple, Charlotte E. Teunissen, Chihiro Sato, Celeste M. Karch

## Abstract

Neuronal dysfunction has been extensively studied as a central feature of neurodegenerative tauopathies. However, across neurodegenerative diseases, there is strong evidence for active involvement of immune cells like microglia in driving disease pathophysiology. Here, we demonstrate that tau mRNA and protein are expressed in microglia in human brains and in human induced pluripotent stem cell (iPSC)-derived microglia like cells (iMGLs). Using iMGLs harboring the *MAPT* IVS10+16 mutation and isogenic controls, we demonstrate that a tau mutation is sufficient to alter microglial transcriptional states. We discovered that *MAPT* IVS10+16 microglia exhibit cytoskeletal abnormalities, stalled phagocytosis, disrupted TREM2/TYROBP networks, and altered metabolism. Additionally, we found that secretory factors from *MAPT* IVS10+16 iMGLs impact neuronal health, reducing synaptic density in neurons. Key features observed *in vitro* were recapitulated in human brain tissue and cerebrospinal fluid from *MAPT* mutations carriers. Together, our findings that *MAPT* IVS10+16 drives cell-intrinsic dysfunction in microglia that impacts neuronal health has major implications for development of therapeutic strategies.

## Introduction

Frontotemporal lobar degeneration (FTLD) encompasses a range of heterogeneous neurodegenerative disorders. Familial FTLD with tau inclusions (FTLD-tau) is characterized by dominantly inherited mutations in the microtubule-associated protein tau (*MAPT*) gene, while common variants in *MAPT* can also play a role in sporadic forms of FTLD-tau ^1, 2^. FTLD-tau is defined by neuropathological hallmarks such as atrophy of the frontal and temporal lobes, abnormal tau protein inclusions, gliosis, and neuronal loss, as well as clinical hallmarks such as behavioral/language difficulties and cognitive dysfunction ^3^. In the brain, *MAPT* is alternatively spliced to produce 6 major isoforms that differ based on N-terminal insertions and the number of repeats in the microtubule binding domain (3 repeats (3R); 4 repeats (4R)). A subset of *MAPT* mutations (e.g. *MAPT* IVS10+16) alter splicing and shift tau isoforms (4R>3R). These mutations mimic a subset of sporadic tauopathies in which 4R tau protein accumulates in brains (e.g., progressive supranuclear palsy and corticobasal degeneration) ^4^.

Tau is a neuronal protein highly concentrated in axons, with minimal presence in other cellular compartments or non-neuronal cells ^5, 6^. Nevertheless, tau-positive inclusions have been documented in astrocytes and oligodendrocytes in human neuropathological studies, including in patients carrying the *MAPT* IVS10+16 mutation ^7–11^. Interestingly, microglial activation has been observed in presymptomatic *MAPT* mutation carriers, even in the absence of protein aggregation and atrophy in the frontotemporal regions ^12, 13^. Furthermore, increased microglial activation has been proposed as a potential biomarker for FTD linked to intronic mutations in *MAPT*, correlating with a more rapid cognitive decline ^14–16^. Despite the evidence of tau inclusions in non-neuronal cells, little work has been done to evaluate *MAPT* mRNA and tau protein expression in glia ^17–19^. This may be due to the prevailing viewpoint that glial inclusions arise from glial cells internalizing tau ladened neurons or capturing tau protein released by neurons ^10, 20, 21^. Microglia, the resident immune cells of the brain, may also play a critical role in tauopathy pathophysiology ^22–25^. Microglia can engulf neurons and synapses burdened with tau ^26–28^. Impaired microglia-mediated clearance of phagocytosed tau can lead to propagation of tau pathology through secretion and promote a vicious cycle that may drive pathological decline ^10, 25, 29, 30^. However, studies directly investigating the potential role of microglia as cell autonomous molecular drivers of tauopathy have been limited. Most studies of the contribution of microglia to tauopathy use animal models that express the human mutant tau transgene in neurons (and not microglia), which does not fully recapitulate human tau expression patterns ^31, 32^. Thus, cell-intrinsic dysfunction of microglia in tauopathies remains largely unknown ^33^.

To begin to understand the contribution of tau to microglia dysfunction in tauopathies, we demonstrated that *MAPT* mRNA and tau protein are expressed in microglia isolated from human brains and in human induced pluripotent stem cell (iPSC)-derived microglia-like cells (iMGLs). We then leveraged iMGLs from *MAPT* IVS10+16 mutation carriers and isogenic controls to discover broad molecular and cellular defects in *MAPT* IVS10+16 iMGLs including dysregulated microglial states, altered cytoskeleton organization, reduced phagocytosis of human myelin and recombinant human tau preformed fibrils, and altered TREM2/TYROBP signaling. *MAPT* IVS10+16 shifts the secretome of iMGLs, which leads to elongated dendrites with reduced synapse density in human neurons. Many of the features we report *in vitro* are maintained in brains and cerebrospinal fluid (CSF) from *MAPT* mutation carriers. Our findings reveal that cell-intrinsic dysfunction in microglia may contribute to neuronal dysfunction and FTLD-tau pathophysiology.

## Results

### FTLD-causing MAPT IVS10+16 mutation causes a shift in microglia transcriptional states

Microglial activation is a key feature of FTLD-tau pathology ^16, 34–36^; however, the cell intrinsic effects of microglia in FTLD-tau pathophysiology remain poorly understood. Tau protein was originally identified as a neuronal microtubule-associated protein specifically localized and enriched in the neuronal axons in the central nervous system (CNS) ^5^. More recently, *MAPT* mRNA expression and tau protein levels have been reported in human iPSC-derived astrocytes ^37^, suggesting that tau may be expressed more broadly in CNS cells than previously thought. To determine whether tau is expressed in microglia *in vivo*, we used a previously reported cohort of 10 human brains in which microglia were isolated and profiled by RNA sequencing ^38, 39^ (**Figure 1A; Supplemental Table 1**). These isolated microglia were enriched for microglia and lacked other contaminating lymphocytes. We found that microglia from all donors expressed *MAPT* (**Figure 1B**), and the overall levels of *MAPT* were robust amongst all protein coding genes captured in the samples (**Figure 1C**). Thus, we demonstrate that human microglia express *MAPT* mRNA *in vivo*.

**Figure 1.**
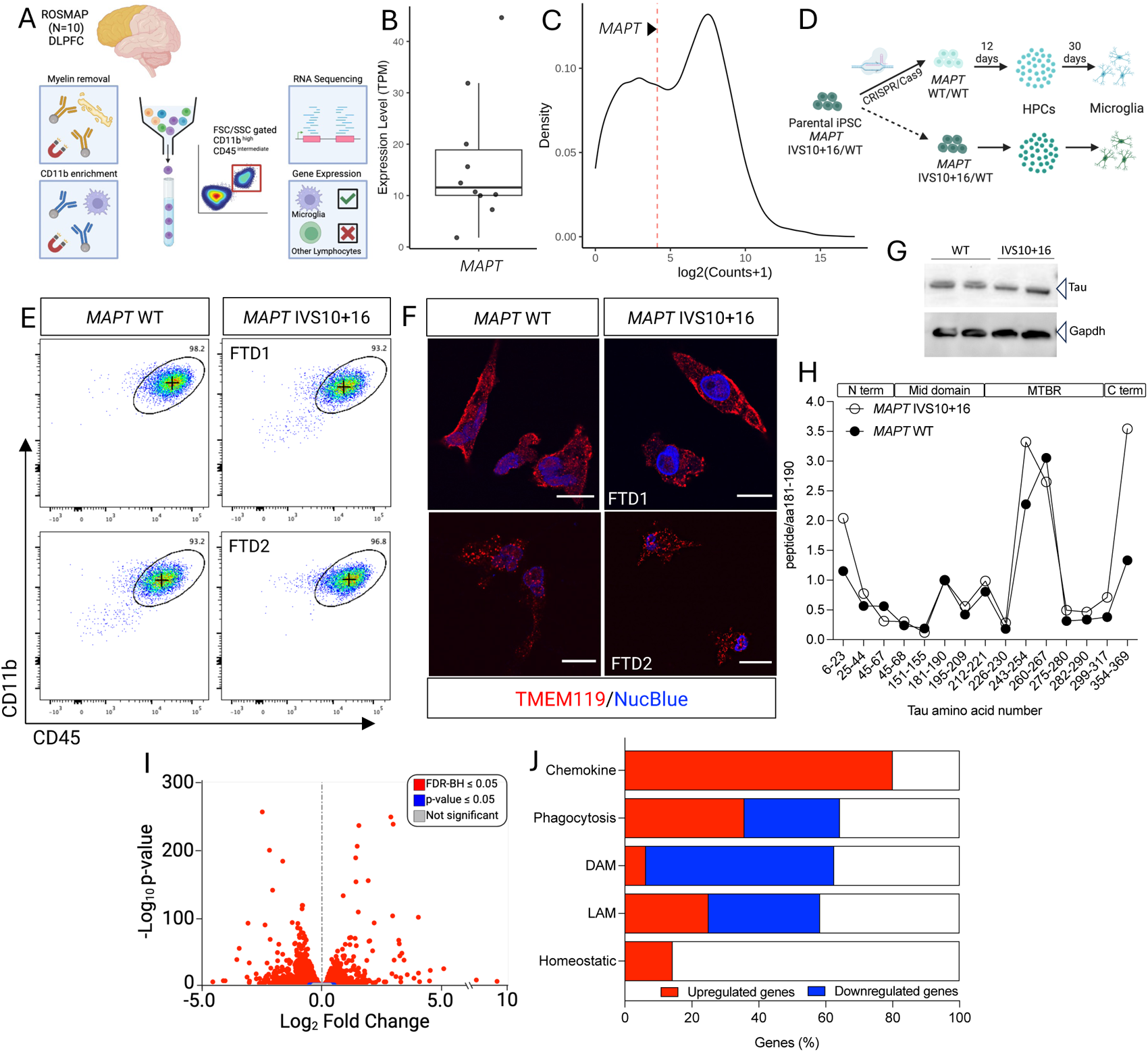
iPSC-derived microglia expressing *MAPT* IVS10+16 reveal shifts in microglia states compared with isogenic controls. A-C. Microglia isolated from human brains and analyzed by bulk RNA sequencing were analyzed for *MAPT* expression (n=10) as previously reported in Olah et al ^39^. A. Workflow for microglia isolation. B. *MAPT* expression (TPM). Each dot represents microglia sampled from an individual brain. C. *MAPT* expression, indicated by the red dashed line, relative to the 19,182 protein coding genes in microglia isolated from human brains. D. Schematic representing how iPSC-derived human microglia-like cells (iMGLs) were generated from hematopoietic progenitor cells (HPCs) differentiated from two independent donors of *MAPT* IVS10+16 carriers: FTD1 and FTD2 and their isogenic controls (4 lines total). E. Dotplots of CD45 and CD11b expression reveals that >95% of the iMGLs are double positive for CD45 and CD11b across donors and genotypes. F. Immunocytochemistry of iMGLs for TMEM119. Scale bar, 12µm. G-H. Whole cell lysates were isolated by detergent extraction and analyzed by immunoblotting for total tau (Tau5) and GAPDH (G) and by immunoprecipitation (Tau1/HJ8.5/HJ8.7 antibodies) and mass spectrometry (H). Peptides monitored by mass spectrometry are plotted relative to the mid-domain peptide at amino acid 181-190 (TPSL). Tau protein domains are annotated above the graph N term (N-terminus), Mid domain, MTBR (microtubule binding region) and C term (C-terminus). Open circles, *MAPT* IVS10+16. Closed circles, *MAPT* WT (isogenic). I. RNAseq and differential gene expression were performed on iMGLs from MAPT IVS10+16 and isogenic controls. Volcano plot. Red dots, FDR-BH ≤ 0.05. Blue dots, p-value ≤ 0.05. Gray dots, not statistically significant. J. Percentage of genes defining specific microglia functions/states: chemokines (n=12 genes), phagocytosis (n= 14 genes), DAM (n= 17 genes), LAM (n=12 genes) and homeostatic (n= 10 genes) were explored in the differential gene expression. Red, significantly upregulated. Blue, significantly down regulated. White, not statistically significant. FDR-BH ≤ 0.05. DAM, disease associated microglia. LAM, lipid-associated myeloid cells.

To understand the microglia cell intrinsic contributions of a mutation in the *MAPT* gene that causes autosomal dominant FTLD-tau, we leveraged human iPSCs from *MAPT* mutation carriers ^40^. iPSCs from two independent donors who are heterozygous carriers of the *MAPT* IVS10+16 mutation (*MAPT* IVS10+16/WT; FTD1 and FTD2) were compared with CRISPR-corrected isogenic controls in which the mutant allele was corrected to WT (*MAPT* WT/WT; **Supplemental Table 1**; **Supplemental Figure 1**; **Figure 1D**). iPSCs were differentiated into iMGLs using a growth factor-based approach through hematopoietic progenitor cells (HPCs) as previously described ^41, 42^ (**Figure 1D**; see Materials and Methods). Using flow cytometry, we demonstrated that iPSC lines, regardless of mutation status, share a similar propensity to form CD45 and CD11b double-positive iMGLs (**Figure 1E**). iMGLs were also similar in their expression of the microglia marker TMEM119 (**Figure 1F**). Thus, the *MAPT* IVS10+16 mutation does not lead to overt defects in the capacity to form mature microglia from iPSCs.

In neurons, *MAPT* IVS10+16 alters tau splicing, leading to elevated 4R tau protein expression ^43^. To determine whether iMGLs express tau and the degree to which *MAPT* IVS10+16 alters tau splicing in iMGLs, we measured *MAPT* mRNA and tau protein. We found that *MAPT* mRNA is expressed in iMGLs (**Supplemental Table 2**) and tau protein is detectable by immunoblotting (**Figure 1G**). Total *MAPT* mRNA and tau protein levels were similar between *MAPT* IVS10+16 and isogenic control iMGLs (**Figure 1G**; **Supplemental Table 2**). To more quantitatively measure tau isoforms, we used immunoprecipitation and mass spectrometry (**Figure 1H**) and discovered that tau isoform expression is similar to patterns we have previously described in human iPSC-derived neurons ^44^, the predominant tau isoform expressed is 3R0N. Additionally, *MAPT* IVS 10+16 iMGLs produced more 4R tau isoforms compared to WT, consistent with human iPSC-derived neurons (**Figure 1H**)^43^. Thus, we demonstrate that iMGLs express the tau mRNA and protein, and *MAPT* IVS10+16 alters tau splicing in iMGLs.

Microglia are heterogenous cells that can exist in diverse, transitory states that are influenced by genetics, environment, development, infection, and disease ^45, 46^. To begin to understand the extent to which the *MAPT* IVS10+16 mutation impacts microglial transcriptional states, we analyzed mutant and isogenic control iMGLs by RNA sequencing (**Figure 1I**; **Supplemental Figure 1; Supplemental Table 2**). Principal component analyses (PCA) revealed that the presence of the *MAPT* IVS10+16 mutation was sufficient to drive global transcriptional signatures (**Supplemental Figure 1E**). To determine whether the *MAPT* IVS10+16 driven transcriptional changes translate to shifts in microglia states, we plotted differential gene expression within each of the key microglial states ^38, 47–51^: chemokine; phagocytosis; disease associated microglia (DAM); lipid associated microglia (LAM); and homeostatic microglia (**Figure 1J**; **Supplemental Table 3**). Chemokines, which play a critical role in chemoattraction, were broadly significantly upregulated in *MAPT* IVS10+16 iMGLs. A number of genes involved in phagocytosis, DAM, and LAM were significantly dysregulated in mutation iMGLs, while homeostatic genes were largely unchanged (**Figure 1J**; **Supplemental Table 3**).

### The cytoskeletal network is altered in iMGLs expressing MAPT IVS10+16

As a microtubule-associated protein, tau has been well characterized for its mechanistic roles in polymerization, stabilization of microtubules, and suppression of microtubule dynamics ^52^. Tau may also serve as a molecular linker that enables interaction between the actin and microtubule cytoskeletal networks to support cellular migration, morphological changes to stimuli, and a host of other cellular functions ^53, 54^. Tau mutations and post-translational modifications have been shown to lead to mis-regulation of microtubule interaction and function ^55–57^. Transcriptomics data revealed a significant upregulation of genes involved in semaphorin signaling and downregulation of genes belonging to the Rho family of GTPases in *MAPT* IVS10+16 iMGLs (**Figure 2A**), both of which have been reported to act as molecular switches in their regulation of actin filament and microtubule networks ^58–60^. To determine whether these cytoskeletal networks are altered in mutant microglia, we performed immunocytochemistry to visualize and characterize the IZl-tubulin (**Figure 2B-C**) and filamentous actin (F-actin) (**Figure 2D-F**) networks in *MAPT* IVS10+16 and WT iMGLs. In *MAPT* WT iMGLs, IZl-tubulin exhibits a radial array pattern originating from the microtubule organizing center (MTOC) (**Figure 2B-C**; **Supplemental Figure 2**). However, in the presence of the mutant allele, IZl-tubulin distribution is shifted to the periphery (**Figure 2B-C**; **Supplemental Figure 2**). Semaphorin signaling and the RhoH GTPase cycle contribute to the disassembly of F-actin ^61, 62^. Consistent with the dysregulation of these pathways in mutant iMGLs, we observed a significant reduction of F-actin (phalloidin) in *MAPT* IVS10+16 iMGLs in comparison to the isogenic WT iMGLs based on immunocytochemistry (**Figures 2D-E**) and flow cytometry (**Figures 2F-G**). Thus, *MAPT* IVS10+16 is sufficient to alter actin and tubulin cytoskeletal networks in microglia.

**Figure 2.**
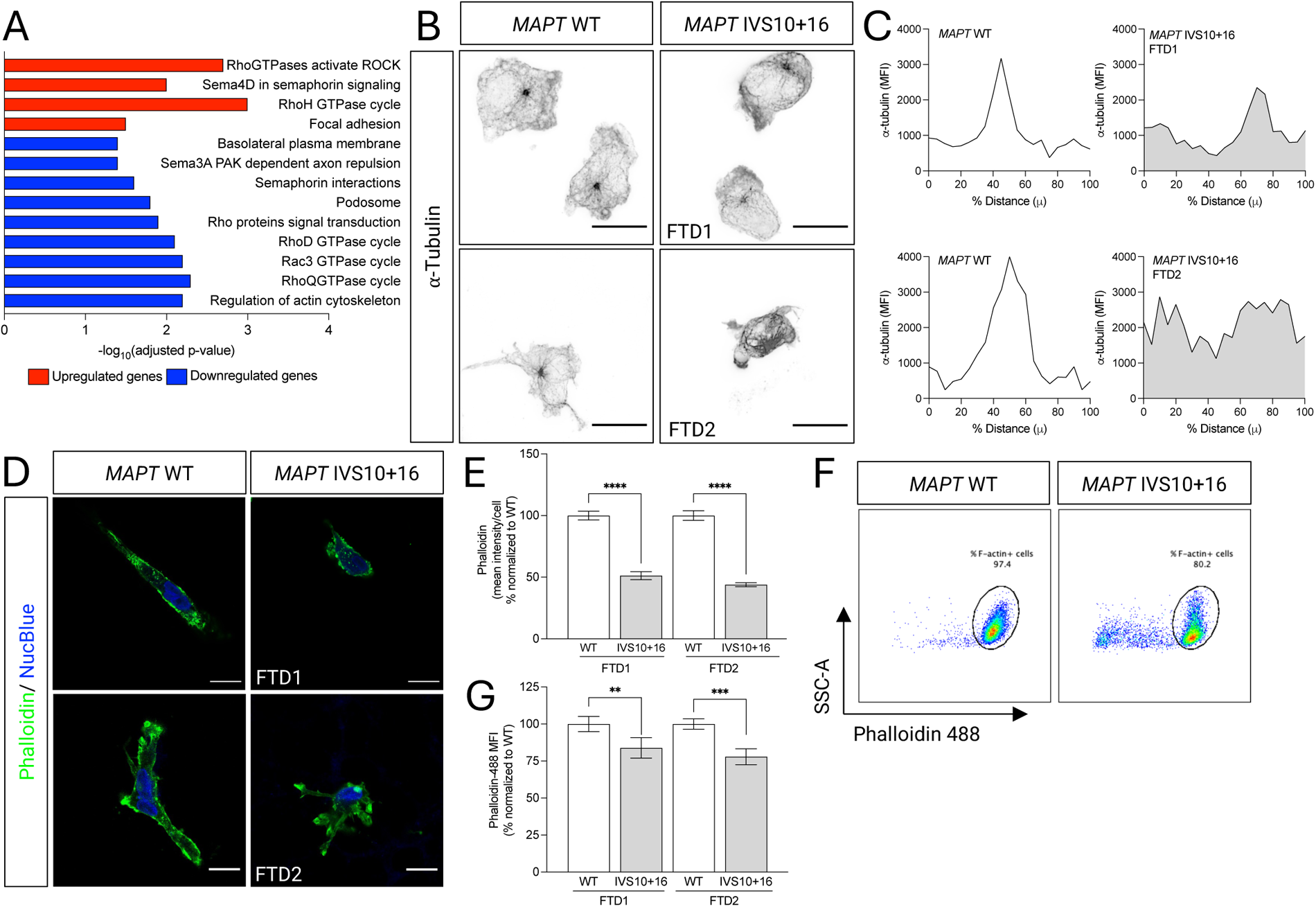
*MAPT* IVS10+16 alters the cytoskeleton in iMGLs. A. Pathway analysis of genes differentially expressed between *MAPT* IVS10+16 and isogenic controls shows an enrichment of genes regulating the cellular cytoskeletal network. Red bars, significantly upregulated genes. Blue bars, significantly downregulated genes. Significance defined as FDR-BH≤0.05. Three biological replicates were included for each donor. B. Immunocytochemistry for IZl-tubulin. Representative images from a Nikon spatial array confocal images acquired at 60x. Scale bar, 12µm. C. Quantification of mean fluorescence intensity (MFI) of IZl-tubulin staining across cellular area using the straight-line tool in ImageJ. MFI was plotted relative to distance across the cell (normalized and expressed as a percentage). D. Immunocytochemistry for filamentous actin (F-actin; phalloidin; green) and nucleus (NucBlue). Scale bar, 12µm. E. Quantification of mean fluorescence intensity of phalloidin within each cell. ****, p<0.0001; *, p<0.05. F. Total intracellular levels of F-actin were quantified by flow cytometry using Phalloidin-488 represented as dot plots reveal the percentage of F-actin positive cells within CD45+CD11b+ iMGLs. Representative dot plots from FTD1 donor pair. G. Quantification of phalloidin (MFI). MFI normalized to WT. Results represent three independent experiments. **, p<0.005; ***, p<0.0005; p-values generated by one-way ANOVA with Sidak’s multiple comparisons test in (E) and (G).

### MAPT IVS10+16 iMGLs exhibit impaired phagocytosis of myelin and tau preformed fibrils

As the CNS resident immune cells, microglia perform constant surveillance functions ^63^. Microglia are required to adapt their cell morphology for rapid migration through narrow spaces in the parenchyma and to phagocytose debris or foreign cargo in their microenvironment ^64^. Efficient microglial phagocytosis relies on downstream signaling cascades that mediate regulation of F-actin networks, which mobilizes around the phagocytic cup to enable target cargo engulfment ^65^. Consistent with our observation that mutant iMGLs exhibited reduced F-actin levels and an altered microtubule network, transcriptomics analyses revealed a significant downregulation of genes involved in phagocytosis and amoeboid-type cell migration, including *MERTK* (**Figure 3A; Supplemental Table 2**). *MAPT* IVS10+16 iMGLs also exhibited a significant upregulation of genes involved in the negative regulation of phagocytosis, suppression of phagosomal maturation, and the prevention of phagosomal-lysosomal fusion (**Figure 3A**).

**Figure 3.**
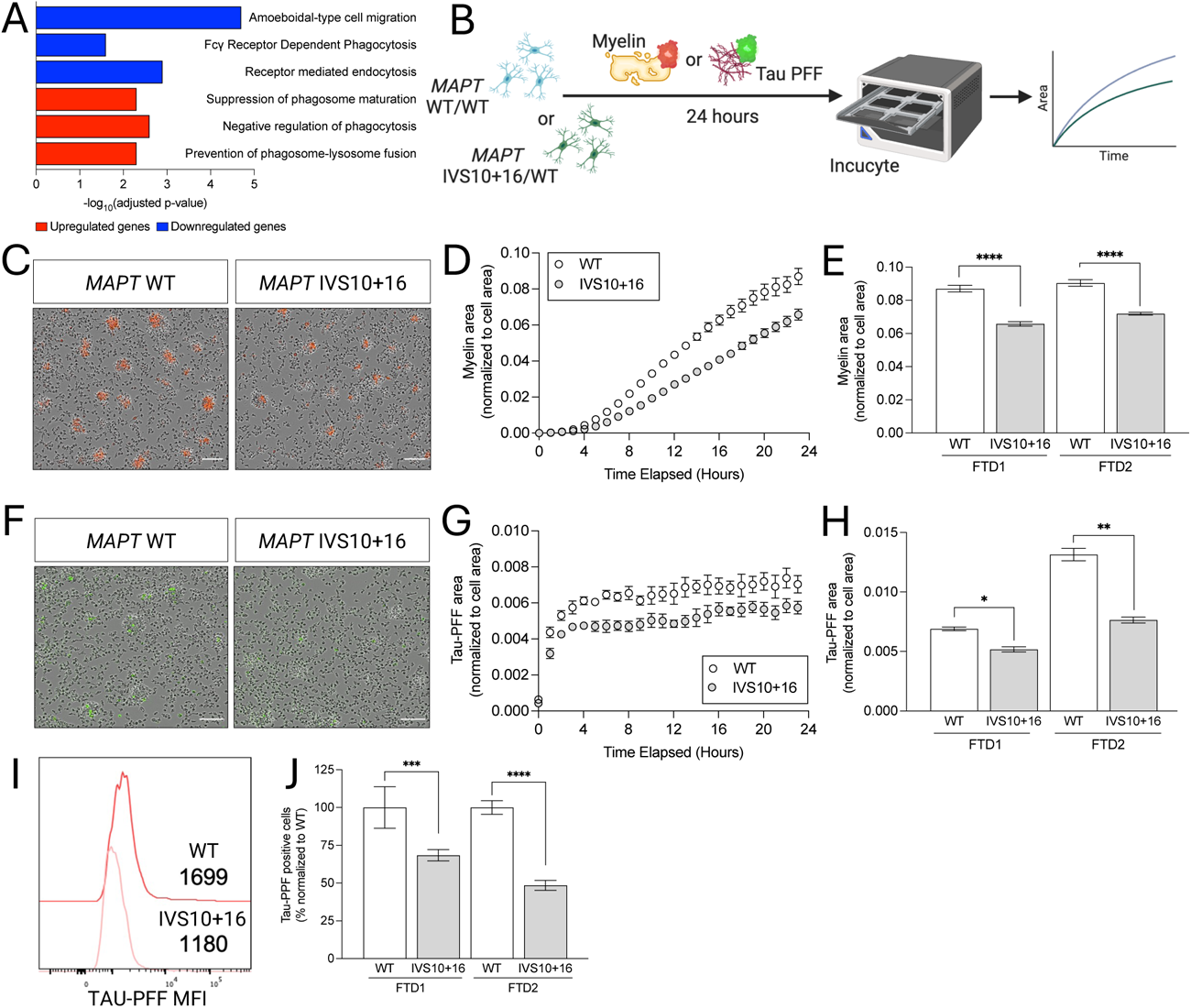
*MAPT* IVS10+16 iMGL exhibit reduced phagocytosis of myelin and tau fibrils. A. Pathway analysis of genes differentially expressed between *MAPT* IVS10+16 and isogenic controls shows an enrichment of genes regulating endocytosis and phagocytosis. Red bars, significantly upregulated genes. Blue bars, significantly downregulated genes. Significance defined as FDR-BH≤0.05. Three biological replicates were included for each donor. B. Phagocytic uptake of human myelin (20µg/ml, C-E) or recombinant human tau preformed fibrils, Tau-PFF (500nM, F-H) analyzed by IncuCyte live cell imaging over 24 hours. C. Representative images 12 hours post myelin treatment. Scale bar, 100µm. D. Representative quantification of myelin area over time. E. Quantification of myelin area at 24 hours post-treatment. ****, p<0.0001. F. Representative images 12 hours post Tau-PFF treatment. Scale bar, 100µm. G. Representative quantification of Tau-PFF area over time. H. Quantification of Tau-PFF area at 12 hours post-treatment. *, p<0.05; **, p<0.005. Data in C-H is representative of 8 replicates per donor line per treatment group. I-J. Tau-PFF uptake was measured by flow cytometry. I. Histogram of Tau-PFF mean fluorescence intensity (MFI) within CD45+CD11b+ cells 24 hours post-treatment. J. Quantification of Tau-PFF MFI. Data is representative of three biological replicates per line. ***, p<0.005; ****, p<0.0001. p-values by two-ANOVA with Sidak’s multiple comparisons test in (E), (H) and (J).

Studies investigating the role of microglia in tauopathies have shown that microglia phagocytose extracellular tau or neurons laden with tau aggregates ^10, 26^. When microglia fail to degrade engulfed tau, they become hypophagocytic, senescence-like, and release tau seeds ^29, 30, 66, 67^. To date, how the expression of mutant tau in microglia can impact these functions has not been well explored. To investigate the direct cell autonomous impact of *MAPT* IVS10+16 on microglial phagocytosis, we challenged iMGLs with physiological (myelin and zymosan A bioparticles) and disease-relevant (tau preformed fibrils (PFF)) substrates and performed hourly live cell imaging for 24 hours (**Figure 3B**). *MAPT* IVS10+16 iMGLs phagocytosed significantly less myelin than isogenic controls (**Figure 3C-E**); however, there was no difference in zymosan A uptake (**Supplemental Figure 3**). Tau PFFs were taken up at a significantly reduced rate in the *MAPT* IVS10+16 iMGLs compared with isogenic controls (**Figure 3F-H**). We further validated the defects in tau uptake in *MAPT* IVS10+16 iMGLs by measuring Tau-PFF fluorescence 24 hours after its addition using flow cytometry (**Figures 3I-J**). Thus, *MAPT* IVS10+16 drives phagocytosis deficits in iMGLs that are substrate specific to dying neurons and tau aggregates.

### TREM2 signaling and metabolic networks are altered in MAPT IVS10+16 iMGLs

TREM2 is a microglia-enriched membrane protein that binds to adapter proteins and signaling mediators like DAP12 (also known as TYROBP) or DAP10 ^68^. This interaction initiates a signaling cascade in microglia that can result in integrin activation, cytoskeleton rearrangement, MAPK signaling, mTOR signaling, calcium mobilization, and energy metabolism ^69, 70^. Transcriptomic analyses revealed a significant downregulation of *TREM2* and TREM2-associated TYROBP network in *MAPT* IVS10+16 iMGLs (**Figure 4A**). Interestingly, we observed a loss of TREM2 staining by immunocytochemistry (**Figure 4B**) and a significant reduction of TREM2 at the cell membrane by flow cytometry (**Figure 4C-D**) in mutant iMGLs.

**Figure 4.**
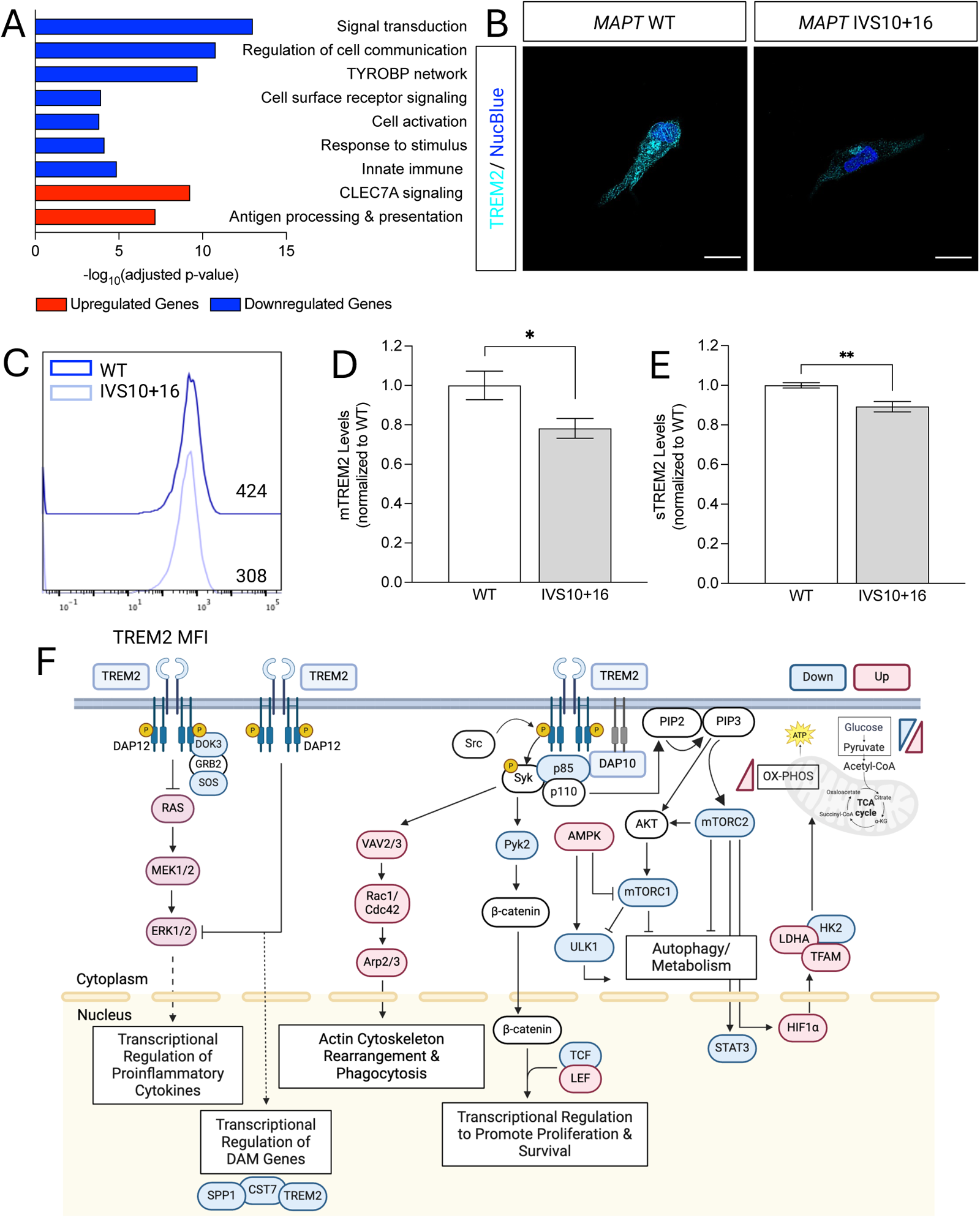
*MAPT* IVS10+16 disrupts TREM2 signaling in iMGLs. Pathway analysis of genes differentially expressed between *MAPT* IVS10+16 and isogenic controls shows an enrichment of genes involved in TREM2 signaling. Red bars, significantly upregulated genes. Blue bars, significantly downregulated genes. Significance defined as FDR-BH≤0.05. Three biological replicates were included for each donor. B. Immunocytochemistry for TREM2 (cyan) and nucleus (NucBlue) Scale bar represents 12µm. C-D. TREM2 on the cell membrane (mTREM2) was measured by flow cytometry. C. Histogram overlay of mTREM2 MFI. D. mTREM2 MFI quantification in both donor pairs. *, p<0.05. E. Soluble TREM2 (sTREM2) levels measured by ELISA in media from both donor pairs. ***, p<0.05; p-values were calculated using an unpaired t-test with Welch’s correction in (D) and (E). F. TREM2 mediates pathways in microglia that regulate actin/cytoskeletal organization, proliferation and survival, cytokines, autophagy, and metabolism. Those genes involved in these pathways that were significantly differentially expressed FDR-BH<0.05 are annotated in red (upregulated in mutant iMGL) or blue (downregulated in mutant iMGL).

Once at the cell surface, TREM2 is cleaved and released as a soluble form (sTREM2) ^71–73^. Increased sTREM2 levels have been equated with increased microglial activity ^74, 75^. In *MAPT* IVS10+16 iMGLs, sTREM2 levels were significantly reduced compared with isogenic controls (**Figure 4E**). Consistent with the downregulation of TREM2 protein and TREM2 signaling pathways, we observed a broad effect on genes and pathways regulated by TREM2 (**Figure 4F**). In addition to altering genes in TREM2-related pathways that contribute to transcriptional regulation of proinflammatory cytokines, transcriptional regulation of DAM genes, actin cytoskeleton rearrangement, and phagocytosis (**Figure 4F**), we also observed a significant reduction in *MTOR* and other endolysosomal genes (**Figure 4F**; **Supplemental Figure 4A**). MTOR is an important regulator of cellular metabolism in microglia ^76–78^. Consistent with these findings, we identified a broad number of genes enriched in glycolysis that were altered in *MAPT* IVS10+16 iMGLs (**Supplemental Figure 4B-C**). *MAPT* IVS10+16 iMGLs exhibited a significant increase in genes involved in mitochondrial oxidative phosphorylation, the mechanism by which cells translate nutrients into energy (**Supplemental Figure 4B-C**).

Together, these findings suggest that the *MAPT* IVS10+16 mutation impacts TREM2 expression and signaling that may ultimately contribute to broad defects in phagocytosis, cytoskeletal organization, endolysosomal function, and metabolism functions in microglia.

### MAPT IVS10+16 iMGL secretome alters neuronal synapses

Here we show that *MAPT* IVS10+16 is sufficient to broadly disrupt microglial transcriptional states and key microglial functions. We identified genes and pathways associated with the adaptive immune response, the innate immune function, metabolism, and lysosomal functions that are dysregulated in mutant microglia (**Supplemental Figure 4D**). To determine the extent to which the genes we detected *in vitro* are reflected in the pathologic processes in FTLD-tau patients, we compared the iMGL transcriptomics with transcriptomics previously generated from the middle temporal gyrus of *MAPT* IVS10+16 mutation carriers and neuropathology free controls (**Supplemental Table 1**) ^79^. We identified 100 genes that were significant differentially expressed in the same direction in mutant iMGLs and FTLD-tau brains (**Supplemental Table 4**). These genes were enriched in pathways associated with adaptive immune activation, NF-κB, inflammasome activation, endolysosomal transport, cell adhesion molecules, and cytoskeletal organization (**Figure 5A**). Interestingly, we also observed a number of the commonly dysregulated genes in pathways involving synaptic assembly and signaling, synaptic vesicle membrane organization, regulated exocytosis, and presynaptic endocytosis (**Figure 5A**).

**Figure 5.**
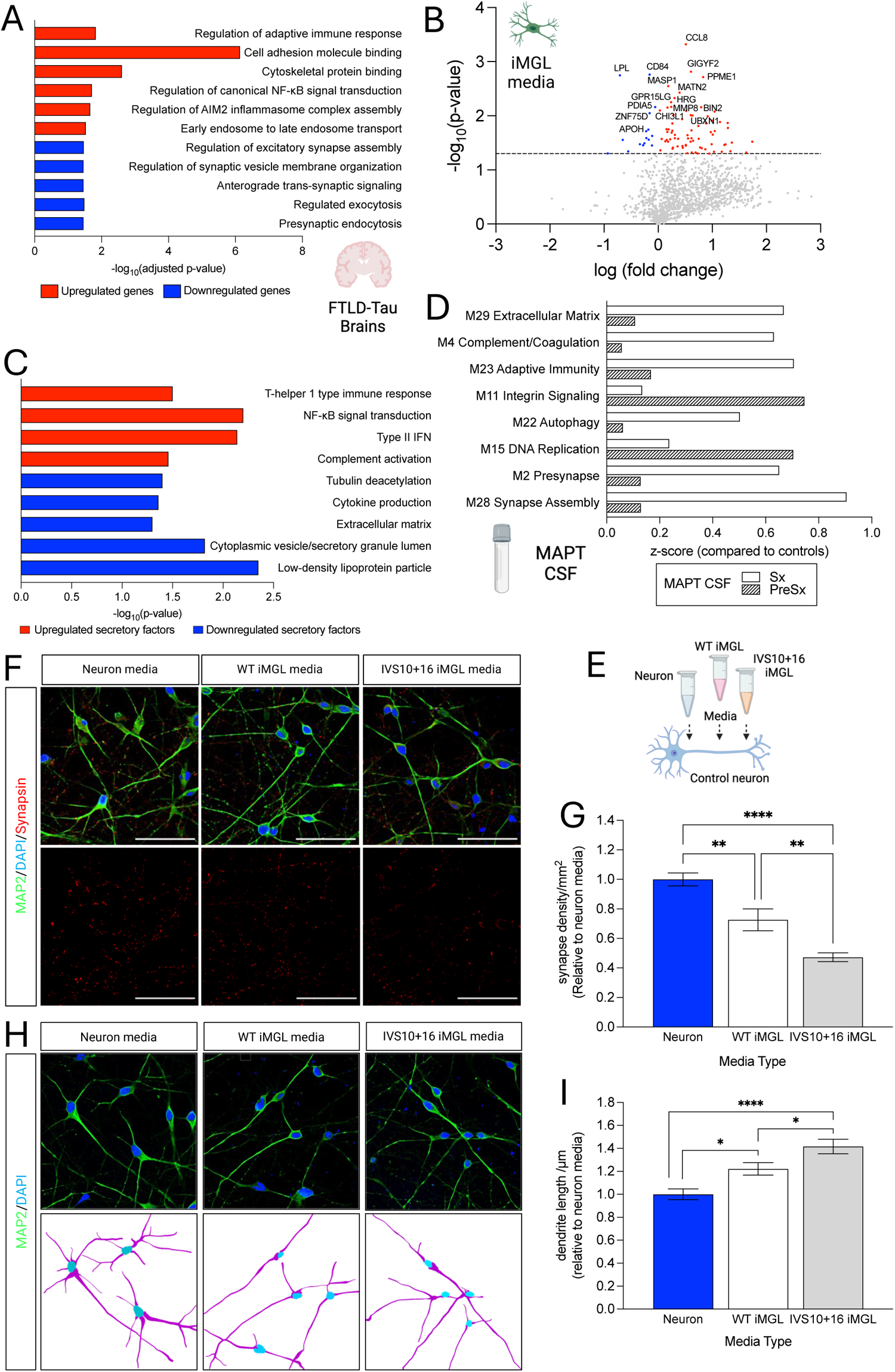
*MAPT* IVS10+16 iMGLs produce secretory factors that alter synapse density. A. Genes shared between *MAPT* IVS10+16 iMGLs and brains of patients with FTLD-tau with *MAPT* mutations (n=100 genes) were enriched in pathways involved in synaptic function and immune response. Genes defined by BH-FDR ≤0.05 in each dataset. B-C. Secretory proteins were monitored using OLINK. B. Volcano plot showing secretory factors altered in comparisons of media from *MAPT* IVS10+16 iMGLs and controls. Three technical replicates were included for each donor pair. Red dots, significantly upregulated (p<0.05). Blue dots, significantly downregulated (p≤0.05). Gray dots, not significant. C. Pathway analysis of differentially expressed secretory factors (p<0.05). Red, upregulated genes. Blue, downregulated genes. D. Proteins in cerebrospinal fluid (CSF) were measured by SomaScan proteomics platform in *MAPT* mutation carriers compared with normal controls. Network analyses revealed several modules that were significantly enriched in CSF from *MAPT* mutation carriers compared with controls. Sx, symptomatic. PreSx, pre-symptomatic. E. Diagram of paradigm. F. Media from control neurons, WT iMGL, or *MAPT* IVS10+16 iMGL was added to iPSC-derived neurons for 7 days. Immunocytochemistry was performed for dendritic marker (MAP2; green), pan-synaptic marker (Synapsin; red) and nucleus (NucBlue). Scale bar represents 50µm. G. Quantification of synaptic density/mm^2^. Data was normalized to neurons cultured in neuronal media. **, p<0.05; ****, p-value<0.0001; one-way ANOVA with Sidak’s multiple comparisons test. H. Immunocytochemistry was performed for dendritic marker (MAP2; green) and nucleus (NucBlue). Scale bar represents 50µm. I. Quantification of dendritic length (μm). Data was normalized to neurons cultured in neuronal media. *, p<0.05; ****, p-value<0.0001; one-way ANOVA with Sidak’s multiple comparisons test.

Microglia secrete many soluble factors such as chemokines, cytokines, and neurotrophic factors that are involved in neuronal homeostasis, synaptic pruning and plasticity, immune responses, and tissue repair in the CNS ^80^. Because of a crucial role for microglia-derived secretory factors in their communication with neurons and other CNS cell types, we next investigated the impact of the *MAPT* IVS10+16 mutation on the microglial secretome. Using a proximity extension assay proteomics platform (OLINK), we assessed the levels of 1360 secretory proteins after QC ^81^. Eighty-nine proteins were differentially expressed in *MAPT* IVS10+16 iMGL media compared with isogenic controls (p-value<0.05, **Figure 5B**; **Supplemental Table 5**). The majority of differentially expressed secretory factors (n=73) were significantly upregulated in *MAPT* IVS10+16 iMGL media and enriched in pathways associated with NF-κB signaling, T cell immune response, type II interferon production, and complement activation (**Figure 5C**). The 16 significantly downregulated secretory factors were enriched in pathways associated with lipid particles, cytokine production, tubulin deacetylation, and secretory granule vesicles (**Figure 5C**). To determine whether these pathways mimic changes occurring in cerebrospinal fluid (CSF) from *MAPT* patients, we examined publicly available aptamer-based proteomics data (SomaScan; 4,138 proteins) from 37 *MAPT* carriers and 39 controls ^82^. Network analyses identified pathways enriched in the extracellular matrix, complement, adaptive immunity, autophagy, and synapse function, among others (**Figure 5D**). These pathways are consistent with our molecular and cellular findings in mutant iMGLs *in vitro* and in the pathways shared in FTLD-tau brains.

Secretory factors derived from microglia conditioned medium have been shown to promote neuronal survival and maturation ^83, 84^ by regulating synaptic responses, density, and network connectivity in primary neurons cultured in such microglia conditioned medium ^85–87^. To determine whether media from *MAPT* IVS10+16 iMGL is sufficient to alter healthy neurons, we cultured iPSC-derived neurons driven by dox-inducible Ngn2, which allowed us to generate a homogenous population of excitatory, glutamatergic neurons. After 7 days in culture, iPSC-derived neurons were cultured in naïve neuronal media or media isolated from *MAPT* IVS10+16 or isogenic WT iMGL for an additional 7 days (**Figure 5E**). Neuronal health and synapses were evaluated by immunocytochemistry for MAP2, a neuronal dendritic marker, and synapsin, a synaptic vesicle marker (**Figure 5F**). Quantification of synapse density/mm^2^ revealed a significant reduction in synapse density in neurons cultured with *MAPT* IVS10+16 iMGL media compared with neurons cultured with WT iMGL media (**Figure 5F-G**). We also observed a decrease in synapse density when comparing the naïve neuronal media-treated cells with the WT iMGL media-treated cells (**Figure 5F-G**). Neurons cultured with iMGL media, regardless of mutation status, had significantly longer dendrites (**Figure 5H-I**). *MAPT* IVS10+16 iMGL media produced neurons with the longest dendrites (**Figure 5H-I**). This is consistent with prior work demonstrating a role of microglia secretory factors in synapse maintenance ^83, 84^. Thus, these findings reveal common dysregulated gene/protein expression in *MAPT* IVS10+16 iMGLs and FTLD-patient tissues (brain and CSF) and implicate microglia-derived soluble factors as potential contributors to neurodegeneration in FTLD.

## Discussion

Microglia have traditionally been viewed as cells that respond and react to increasing tau aggregates and neuronal damage in tauopathies. Here, we shed light on the cell intrinsic contributions of microglia expressing a FTLD-causing *MAPT* mutation to disease pathogenesis. We developed a stem-cell-based model to uncover the cell-autonomous molecular effects of *MAPT* mutations specifically in human microglia. *MAPT* IVS10+16 leads to a shift in microglial transcriptional states. We demonstrate that *MAPT* IVS10+16 causes alterations in cytoskeletal arrangement, defects in phagocytosis of myelin and tau PFFs, and reduced TREM2 signaling. Conditioned media from *MAPT* IVS10+16 iMGLs is sufficient to alter neuronal synapse density and dendritic length. Together, these findings have important implications for our understanding of the early involvement of microglia in the pathogenesis of tauopathies.

Here, we report that *MAPT* mRNA and tau protein are expressed in human iMGLs. Notably, mass spectrometry analysis from a pure population of mature iMGLs revealed full length tau protein, consistent with prior reports in iPSC-derived neurons and astrocytes ^37, 43^. In tauopathies, microglia have been shown to play an important role in disease progression. Microglia acquire a hypofunctional, senescence-like and paracrine-mediated state upon phagocytosis of neurons burdened with insoluble tau aggregates. Microglia may go on to serve as vectors of tau spread via exosomes due to an inability to break down disease-related tau ^10, 22, 23, 25, 29, 66, 67, 88–90^. Additionally, microglia-derived secretory factors and their proinflammatory state may initiate tau aggregation in neurons in tauopathy mouse models ^88, 89, 91–93^. However, these studies have relied on mouse models that do not express mutant human tau in microglia and thus, our knowledge of the role of mutant tau within microglia as a driver of disease and the presence of cell-autonomous defects in microglia has remained limited until now.

We show that the *MAPT* IVS10+16 mutation induces changes in the cytoskeleton of microglia, characterized by the redistribution of the tubulin network towards the cell periphery and a decrease in F-actin levels. Tau is associated with the regulation of cytoskeletal organization and dynamics, acting as a connector between the actin and tubulin cytoskeleton during processes like axon outgrowth and growth cone turning in cortical neurons ^52, 53, 55, 94^. Microtubule remodeling is critical for microglial activation ^95, 96^. Interestingly, COS-7 fibroblasts expressing a phosphomimetic form of tau (Y18E) also undergo microtubule redistribution, affecting endosomal and lysosomal motility ^97^. Actin dynamics are crucial for microglial functions such as phagocytosis ^98, 99^, suggesting that the reduced phagocytic capacity of *MAPT* IVS10+16 iMGLs could be linked to lower F-actin levels in these cells. Thus, a worthwhile focus of future studies would be on elucidating the mechanisms by which tau influences cytoskeletal network remodeling in microglia and its impact on microglial phagocytosis.

Our discovery that *MAPT* IVS10+16 leads to downregulation of the TREM2-TYROBP signaling network and a decrease in TREM2 expression at the gene, membrane, protein, and secreted levels suggest tau impacts a major microglia signaling molecule implicated in FTLD-tau and secondary tauopathies such as AD. TREM2 signaling has been shown to play a key role in connecting several microglial functions such as cell survival, proliferation, phagocytosis, endolysosomal pathways, metabolism, and inflammation ^68, 70, 100^. Homozygous mutations in the gene encoding *TREM2*, an activating receptor exclusively expressed in microglia in the CNS, are clinically associated with FTD-like syndrome ^101^. TREM2 plays a crucial role in microglial phagocytic function ^23, 98, 100^. TREM2 missense mutations linked to FTLD and FTLD-like syndrome lead to impaired TREM2 trafficking to the cell surface and reduced microglial phagocytosis, highlighting a direct association between mature membrane-bound TREM2 levels and microglial phagocytic activity ^102^. Studies leveraging *TREM2* loss-of-function mutations in mouse models of tauopathy point to a role for TREM2 signaling in the regulation of amyloid accumulation, tau pathology, and neuroinflammation in Alzheimer’s disease. ^23, 103^. However, whether TREM2 plays a neuroprotective or neurotoxic role in tau pathology may depend on the disease stage in animal models ^104–107^.

Among the many roles of TREM2 in microglia, mounting evidence points to a role in regulating the endolysosomal network and sustaining microglial metabolic balance through mTOR signaling ^77, 78^. Previously, we have shown that *TREM2* loss-of-function mutation in iMGLs leads to lysosomal dysregulation ^77^. Consistent with our prior work, transcriptomics analyses in *MAPT* IVS10+16 iMGLs revealed a downregulation of genes involved in the late endosomal/lysosomal processes. Interestingly, we also observed downregulation of *MTOR*, a sensor of cellular energy and homeostasis, in *MAPT* IVS10+16 iMGLs. TREM2-dependent mTOR signaling is crucial to sustain biosynthetic and energy metabolism in microglia ^78^ and consistent with this, we observed a dysregulation of genes involved in glycolysis and a selective upregulation of genes involved in mitochondrial oxidative phosphorylation in *MAPT* IVS10+16 iMGLs. Maintenance of lysosomal homeostasis is crucial to controlling microglial mTOR levels and initiating immune responses to tau pathology in the tau-P301S tauopathy mouse model ^108^. Microglia activation also induces mitochondrial alterations and initiates a metabolic shift from oxidative phosphorylation to glycolysis ^109^. iMGLs carrying hypofunctional *TREM2* risk variants (e.g. R47H) exhibit deficiencies in their ability to transition from oxidative phosphorylation (OXPHOS) to glycolysis, along with reduced mitochondrial respiration rates ^110^. Our findings point to a role for *MAPT* IVS10+16 in suppression of TREM2 and associated gene networks that regulate cell survival, proliferation, phagocytosis, endolysosomal pathways, metabolism, and inflammation. Further studies are needed to understand the molecular mechanisms involved and to explore whether activating TREM2 signaling will rescue the observed defects in phagocytosis, cytoskeletal network, and metabolism in the *MAPT* IVS10+16 iMGLs.

Here, we describe broad alterations in the *MAPT* IVS10+16 iMGL secretome. Both actin and microtubule dynamics are involved in microglial secretory function ^95, 99^. *MAPT* IVS10+16 iMGLs exhibit less secreted lipoprotein lipase (LPL), a protein associated with microglial phagocytosis ^111^. Knockdown of microglia specific *Lpl* in a mouse model of obesity, resulted in overall decreased microglial reactivity, phagocytic ability, mitochondrial disturbances, and an inability to transition to an adaptive immunometabolic state ^112^. Microglia and complement signaling play crucial roles in synapse elimination during developmental pruning and disease, sometimes involving aberrant phagocytosis of synapses and viable neurons ^26, 113–115^.

Interestingly, we found that neurons treated with *MAPT* IVS10+16 conditioned microglial media had fewer synapses. This could be driven by a number of secretory factors that are altered in the *MAPT* IVS10+16 microglia. MASP1, an activator of the lectin pathway of complement proteins, is upregulated in *MAPT* IVS10+16 iMGLs secretome. MASP1 has been implicated in loss of presynaptic inputs in axotomized spinal motoneurons following peripheral nerve injury ^116^. While complement-mediated synapse elimination has been observed primarily through microglial contact-dependent phagocytosis of synaptic terminals ^117^, several studies have demonstrated that microglia can also modulate neuronal synaptic density and network connectivity through soluble secretory factors ^85–87^. Microglial activation and synaptic loss occur prior to the accumulations of fibrillary tau tangles in the P301S tauopathy mouse model ^118^. Microglia-derived secretory factors can trigger tau pathology in neurons ^30, 92^. Thus, our finding that synaptic density is altered when neurons are exposed to *MAPT* IVS10+16 conditioned media suggests a pivotal role for microglia-mediated disease mechanisms in tauopathies. Further work is required to understand whether the secreted proteins that are changed due to *MAPT* IVS10+16 may serve as potential new biomarkers of microglial dysfunction in tauopathies.

In this study, we have developed a stem-cell-based model to uncover the cell-autonomous molecular effects of *MAPT* mutations specifically in human microglia. iPSC-based microglial modeling is a powerful cellular platform that captures endogenous human tau expression to explore the molecular, cellular, and disease-relevant phenotypes resulting from FTLD-causing *MAPT* genetic mutations. By studying these changes in human microglia, we can better identify their contributions to tauopathies and other neurodegenerative diseases.

### Experimental Procedures

#### Patient consent

The informed consent was approved by the Washington University School of Medicine Institutional Review Board and Ethics Committee (IRB 201104178 and 201306108). The University of California San Francisco Institutional Review Board approved the operating protocols of the UCSF Neurodegenerative Disease Brain Bank (from which brain tissues were obtained). Participants or their surrogates provided consent for autopsy, in keeping with the guidelines put forth in the Declaration of Helsinki, by signing the hospital’s autopsy form. If the participant had not provided future consent before death, the DPOA or next of kin provided it after death. All data were analyzed anonymously.

#### Induced pluripotent stem cell (iPSC)

Human iPSC lines derived from dermal fibroblasts were previously described and characterized using standard methods ^79, 119^. *MAPT* IVS10+16 mutation carrying GIH36C2 (FTD1) and GIH178C1 (FTD2), and their corresponding CRISPR-Cas9 corrected isogenic control lines, GIH36C2Δ1D01 and GIH178C1Δ2A10, were used in this study and donor-specific information are provided in **Supplemental Table 1**. Human iPSCs were cultured in mTesR1 on Matrigel-coated tissue culture-treated plates using Accutase™ (ThermoFisher Scientific, Cat#: NC9464543) for dissociation during routine passaging. Each line was analyzed for pluripotency markers by immunocytochemistry (ICC) and quantitative PCR (qPCR); for spontaneous differentiation into the three germ layers by ICC and qPCR; and for chromosomal abnormalities by karyotyping. Mutation status was confirmed in iPSC by Sanger sequencing. Karyotyping and Sanger sequencing to verify mutation status were performed every 20 passages. iPSCs were maintained with less than 5% spontaneous differentiation.

#### Generation of hematopoietic progenitor cells from iPSCs

Human iPSCs were differentiated into microglia using a two-step protocol modified from previous reports ^120^. To generate hematopoietic progenitor cells (HPCs), a STEMdiff Hematopoietic kit (STEMCELL Technologies, Cat#: 05310) was used following manufacturer’s instructions as previously reported ^41, 42, 77^. Briefly, iPSCs were detached with ReLeSR™ (STEMCELL Technologies, Cat#: 05872) and passaged in mTeSR1 supplemented with Rock inhibitor, Y-27632 Dihydrochloride (STEMCELL Technologies, Cat #72302) to achieve a density of 45-80 aggregates/well. On day 0, cells were transferred to Medium A from the STEMdiff Hematopoietic Kit to pattern iPSCs towards mesoderm. On day 3, mesodermal cells were exposed to Medium B to further promote differentiation into HPCs, and cells remained in Medium B for 10 additional days. After 12 days in culture, HPCs were collected for fluorescence activated cell sorting (FACS) to enrich for CD43+ CD34+ HPCs.

#### FACS for HPC generation and banking

For sorting of CD43+ CD34+ HPCs, HPCs were collected from the floating population with a serological pipette and from the adherent population after incubation with Accutase at 37°C for 20 mins. Cells were filtered through 70 μm filters to remove large clumps, then washed with FACS buffer (2% FBS in 1x Dulbecco’s Phosphate Buffered Saline (DPBS), modified without calcium chloride and magnesium chloride, sterile-filtered, Sigma-Aldrich D8537; 300g at 4°C for 5 mins), then immunolabeled in the dark for 20 mins at 4°C in FACS buffer containing the following antibodies: CD43-APC (Biolegend, Cat#: 343206, Clone: 10G7), CD34-FITC (Biolegend, Cat#: 343504, Clone: 581), CD45-Alexa 700 (Biolegend Cat#: 304024; Clone: H130). After immunolabeling, HPCs were washed once with FACS buffer and then resuspended in ∼300 µL FACS buffer and filtered through 5 mL polystyrene round-bottom tube with cell-strainer cap (Falcon, Cat#: 352235) and sorted with a Becton Dickinson FACSAria II cell sorter. All steps were performed on ice and using pre-chilled reagents and a refrigerated centrifuge. After sorting, HPCs were spun down at 300g at 4°C for 10 mins and the supernatant was aspirated. Cell counts were determined using Invitrogen Countess 3 automated cell counter and the cell pellet was then frozen in 1mL CryoStor® CS10 (STEMCELL Technologies, Cat#: 07930) for every 1x10^6^ HPCs and stored in liquid nitrogen until use.

#### Differentiation of HPCs into microglia-like cells (iMGLs)

For iMGL differentiation, HPCs were thawed into 5% FBS in DMEM/F12 and centrifuged at 300g for 6 mins at room temperature (RT). The resulting cell pellet was resuspended in iPSC-microglia medium comprised of DMEM/F12 (Sigma-Aldrich, cat#: D8437), 2X insulin-transferrin-selenite (Gibco, cat#: 41400-045), B-27™ Supplement (50X, 1: 25, Life Technologies, cat#: 17504-044), N-2 Supplement (100X, 1:200, ThermoFisher Scientific, cat#: 17502001), glutamax (1X, ThermoFisher Scientific, cat#: 35050061), 1X non-essential amino acids (Thermo Scientific, cat#: 11140050), 400 mM monothioglycerol (Sigma Aldrich, cat#: M1753), 5 µg/mL human insulin (recombinant, Sigma Aldrich, cat#: I2643), and freshly supplemented maintenance cytokines (100 ng/mL IL-34 [PeproTech, Cat#: 200-34], 50 ng/mL TGFβ1 [PeproTech, Cat#: 100-21], and 25 ng/mL M-CSF [Peprotech, Cat #:300-25]). Cells were then maintained on Matrigel coated 6-well plates. On days 6 and 12 post-HPC generation, iMGLs were split based on cell density per well. Floating and adherent cells from each well were evenly transferred from three wells into one new well to refresh Matrigel and promote proliferation during this phase. On day 25 post-HPC generation, iMGLs were replated onto Matrigel-coated tissue culture in microglia media supplemented with maturation cytokines (100 ng/mL CD200 [Novoprotein, Cat#: C311] and 100 ng/mL CX3CL1 [Peprotech, Cat#: 300-31]) ^77^. All experiments described in this study were performed in mature iMGLs harvested on day 28 post-HPC generation.

#### Assessment of iMGL cell surface marker expression by flow cytometry

To evaluate cell surface markers, iMGLs (28 days post-HPC generation; 100,000 cells) were treated with Zombie Aqua™ dye (Fixable Viability kit, BioLegend, Cat# 423102) for 20 mins in PBS (1:1000) followed by a wash with FACS buffer. iMGLs were incubated for 10 min on ice with anti-CD16/CD32 to block Fc receptors (1:50; Miltenyi Biotec, Cat #:120-000-442). Then, iMGLs were stained with CD11b-PE/Cy7 (Cat#: 101216; Clone: M1/70), CD45-Alexa Fluor 700 (Cat#: 304024; Clone: H130), TREM2-APC (R&D, FAB17291A) for 20 min at 4°C in the dark followed by washing with FACS buffer. Cell surface markers were acquired on a BD X-20 analyzer. The resulting data were analyzed with FlowJo software (FlowJo 10.8.1).

#### Immunoblotting

iMGLs were lysed in 1x RIPA lysis buffer (150mM NaCl, 1% NP-40, 0.5% deoxycholate, 0.1% SDS, 50mM Tris pH-7.4) containing protease inhibitor cocktail (1:500, Sigma, Cat#: P8340) and phosphatase inhibitor (PhosSTOP™, Roche, Cat#: 04906837001) and incubated on ice for 20 mins. Lysates were then centrifuged at 14000g for 20 minutes at 4°C, and the supernatant was isolated for subsequent analyses. To evaluate total protein concentration, a BCA assay (Pierce™ BCA Protein Assay Kits ThermoScientific, Cat#: 23225) was performed. SDS-PAGE was performed using 20 µg of iMGL cell lysates mixed with 4LJ×LJLaemmli sample buffer (Bio-Rad, Cat#: 161-0747) and 10% β-mercaptoethanol and heated at 95 °C for 10 min. A 4–12% bis–tris gel (Nupage) was used. Proteins were transferred to PVDF membrane and blocked for 1 hr at RT in 5% milk in phosphate buffered saline with 0.1% Tween 20 (PBS-T). Membranes were probed with the mouse anti-Tau5 (a generous gift from Binder lab) and GAPDH (1:500, Thermo Fisher Scientific, Cat# MA5-15738, RRID: AB_10977387) overnight at 4°C. Membranes were subsequently washed and incubated in affiniPure Goat anti-mouse HRP (1:2000, Jackson Immuno Research Labs, Cat# 115-035-174, RRID: AB_2338512) for 1 hr at RT, washed, and developed using SuperSignal West Pico PLUS (ThermoScientific, Cat#: 34580) on a Bio-Rad Chemidoc Imaging System.

#### Immunoprecipitation and Mass Spectrometry Analyses

iMGLs were homogenized in 300 µL 50 mM PBS containing 1x Protease inhibitors (Roche) for 10 mins. Lysates were sonicated on ice and centrifugated at 10,000 x g for 20 mins at 4°C to isolate cell lysate. Total protein concentration was determined using the BCA assay method. Tau was then immunoprecipitated using a mixture of Tau1, HJ8.5, and HJ8.7 antibodies, following a previously described protocol with adjustments ^121–123^. For each sample, 2.5 ng of 15N labeled recombinant 2N4R tau isoform (0.5 ng/µL, a gift from Dr. Guy Lippens, France) was added to 30 µg total protein. Soluble tau was immunoprecipitated in a solution containing detergent (1% NP-40), 5 mM guanidine hydrochloride, and protease inhibitors (Roche), using a tau antibody-conjugated sepharose bead cocktail (50% slurry containing 3 µg antibody/mg beads). The immunoprecipitation was conducted by rotating the samples overnight at 4 °C. After incubation, sepharose beads were washed with 25 mM TEABC buffer. Immobilized proteins were digested on the beads overnight (16-18 hrs) using 0.4 µg trypsin at 37 °C. The digested peptides were diluted with 300 µL of a 3% oxidation solution (Formic acid + H_2_O_2_) and incubated overnight at 4 °C. The digested and oxidized samples (soluble tau) were desalted using the Oasis µElution HLB plate (Waters^TM^ Corp) following the manufacturer’s protocol. The eluent was lyophilized and reconstituted with 25 µL of 2% ACN, 0.1% FA in water prior to mass spectrometry analysis on Vanquish Neo UHPLC (Thermo Scientific) coupled to Orbitrap Exploris 480. Trypsinized tau peptides were quantified by comparing with the corresponding isotopomer signals from the 15N internal standard using Skyline software (version 20.2, MacCoss Lab, Department of Genome Sciences, University of Washington).

Tau peptides as prepared above were loaded directly onto an HSS T3 75 μm × 100 μm, 1.8 μm C18 column (Waters) at a temperature set to 65 °C using a Vanquish Neo UHPLC (Thermo Fischer Scientific, San Jose, CA, USA). Peptides were separated with a flow rate of 0.4 µL/min using buffer A (0.1% FA in water) and buffer B (0.1% FA in acetonitrile). Tau peptides eluted from the column with a gradient of 4%-8% buffer B for 10 min, followed by 8%-20% buffer B for another 8 min, ramping up to 95% buffer B in the next 2 min, and column cleaning for an additional 2 min. The Thermo Orbitrap Exploris 480 was equipped with a Nanospray Flex electrospray ion source (Thermo Fisher Scientific, San Jose, CA, USA) and operated in positive ion mode. Peptide ions sprayed from a 10 μm SilicaTip emitter (New Objective, Woburn, MA, USA) into the ion source (spray voltage = 2200 V) were targeted and isolated in the quadrupole. Isolated ions were fragmented by HCD, and ion fragments were detected in the Orbitrap (resolutions of 15000, 30000, and 60000 for N-terminal, mid-domain, and MTBR tau peptides, mass range of 150–1,500 m/z).

#### RNA-sequencing

RNA was extracted from iMGLs using RNeasy Mini Kit (Qiagen, Cat#: 74104). Total RNA integrity was determined using Agilent Bioanalyzer or 4200 Tapestation. Library preparation was performed using Ribo-ZERO kits (Illumina-EpiCentre). RNA was sequenced on an Illumina NovaSeq-6000 using paired end reads extending 150 bases. RNA-seq reads were then aligned and quantitated to the Ensembl release 101 primary assembly with an Illumina DRAGEN Bio-IT on-premises server running version 3.9.3-8 software.

#### Principal component and differential expression analyses

Transcriptome analyses were based on 12,272 protein-coding genes. These genes were expressed in ≥ 50% of the biological replicates analyzed in this research and with one or more counts per million (CPM). Principal component analyses (PCA) were calculated using a subset of the 500 most variable genes based on regularized-logarithm transformation (rlog) counts. PCA and Volcano plots were created using ggplot2 ^124^. Differential gene expression was performed using the DESeq2 for R package 4.3.1 ^125^. Unadjusted p-value (≤ 0.05) and Benjamini-Hochberg adjusted p-value (FDR≤0.05) corrections were applied. Genes passing the FDR≤0.05 threshold were analyzed by EnrichR and ToppGene ^126, 127^.

#### Human Brain RNA-sequencing dataset

The bulk RNA-seq dataset for FACS-isolated microglia from ROSMAP was downloaded from the AMP-AD Knowledge Portal and processed as previously described ^38^. Briefly, microglia were collected from postmortem human brains ^38^. Gray matter was mechanically homogenized followed by myelin depletion with antibody-based beads ^38^. The resulting fraction was then enriched for CD11b positive cells using antibody-based beads, and cells were isolated by FACS using CD11b and CD45 ^38^. These isolated microglia were demonstrated to be free of CD45 high macrophages, infiltrating monocytes, and other potentially contaminating cells from the blood. RNA-seq of the FACS-isolated microglia demonstrated high expression of typical microglial genes (*CX3CR1*, *TREM2*, *P2RY12*, *GPR34*, *TMEM119*) and an absence of markers for other blood cells (*GYPA*, *ITGA2B*, *GP1BA*, *SELP*, *CD3D*, *CD3E*, *CD3G*, *NCAM1*, *CD19*, *MS4A1*, *SDC1*, *CCR2*, *ENPP3*, *CD34*, *MCAM*). Ten samples, AD (N = 4) and healthy control (N = 6), were included in the analysis. The sample size, brain region, age, sex, postmortem interval (PMI), and disease status are summarized in **Supplemental Table 1.**

To determine whether the differentially expressed genes in the iMGLs capture molecular processes that occur in primary tauopathies, we analyzed gene expression in human brain RNA-seq datasets (**Supplemental Table 1**) from the middle temporal gyrus from *MAPT* IVS10 + 16 mutation carriers (2 samples) and healthy controls (3 samples) ^79^. Differential expression analyses were performed including sex, age-at-death, RNA integrity number (RIN), and brain tissue source as covariates.

#### Immunocytochemistry

iMGL (50,000 cells per well) were seeded onto 8-well chamber slides (Millicell EZ SLIDE, Cat#: PEZGS0816) on day 25 post HPC generation in iMGL basal medium with maintenance and maturation cytokines. iMGLs were fed on alternate days until 28 days post HPC generation. Cells were washed one time with 1x DPBS and fixed with paraformaldehyde (4% w/v) for 20 min at RT followed by three washes with 1x DPBS. Cells were treated with blocking buffer (1x DPBS with 0.3% Triton X-100 and 3% BSA) for 1 hr at RT followed by permeabilization in 0.5% Triton X-100 in 1x DPBS for 15 mins. Primary antibodies (see below) were then added at specified dilutions in blocking buffer and placed at 4°C overnight. Cells were then washed 3 times with 1x DPBS for 5 min each wash then stained with donkey Alexa Fluor conjugated secondary antibodies (Invitrogen,1:500) in blocking buffer for 2 hrs at RT in the dark. After secondary antibody staining, cells were washed three times with 1x DPBS PBS and cover slipped with Fluoromount-G™ (ThermoFisher).

Primary antibodies were used at the following dilutions in this study: rabbit anti-TMEM119 (1:100, Abcam, Cat#: ab185333), goat anti-TREM2 (1:100, R&D, Cat#: AF1828), rabbit IZl-tubulin (1:100, Abcam, Cat#: ab18251) and Alexa Fluor™ Plus 647 Phalloidin (1:400, Invitrogen, Cat#: A30107). Nuclei were stained with NucBlue™ Fixed Cell ReadyProbes™ Reagent (DAPI; ThermoFisher Scientific, Cat# R37606). Images were acquired in the Nikon AX-R with NSPARC (Nikon’s Spatial Array Confocal detector), a laser scanning confocal microscope. Images were acquired using a 60x, 4-7x zoomed Z-stack images were acquired in the AX NSPARC mode. Image analyses were performed in Fiji. Representative images shown in figures are projected into max intensity z plane and scale bar is as indicated in respective figure legends. The number of cells analyzed for each staining are as indicated in figure legends. Six to eight confocal images acquired at 60x across two independent wells of a 8-well chamber slide were analyzed for quantification of IZl-tubulin quantification shown in Figure 2C. Three to four independent wells with five to thirty cells per well were analyzed for F-actin quantification shown in Figure 2E.

#### Phagocytosis assays

iMGLs (50,000 cells) were seeded into Matrigel-coated flat bottom 96-well plate on day 25 post-HPC generation in iMGL basal medium with maintenance and maturation cytokines and fed on alternate days until day 28. On day 28 post-HPC generation, iMGLs were treated with phagocytic substrates such as human myelin (20 µg/ml, prepared as previously reported in ^77^), tau preformed fibrils (Human Recombinant Tau-441 (2N4R) P301S mutant protein pre-formed fibrils ATTO 488, TAU-PFF, StressqMarq, Cat# SPR-329-A488, 500nM), or Alexa Fluor™ 488 conjugated Zymosan A (S. cerevisiae) BioParticles™ (ThermoFisher Scientific, Cat#Z23373, 0.5 µg/ml). Tau-PFF was prepared for treatment by sonication in water bath at 20A for 3 cycles of 30 seconds on and 30 seconds off. Imaging was performed hourly for 24 hrs following substrate addition using an Incucyte S3 Live Cell Analysis Instrument (Sartorius). Eight biological replicates were used in each experiment. Data analyses were performed using the IncuCyte S3 software v2021A and bar graphs to represent specific time-points were generated in GraphPad Prism 10 (version 10.0.3).

#### Tau uptake - flow cytometry

On day 25 post-HPC generation, 100,000 iMGLs were seeded in Matrigel-coated 24-well tissue culture plates in iMGL basal medium with maintenance and maturation cytokines and fed on alternate days until day 28. iMGLs were either treated with PBS vehicle or 50 nM TAU-PFF (sonicated as described above) on day 28 for 24 hrs. Cells were then harvested on ice from the 24-well plates, washed thrice with 1x DPBS, and replated into U-bottom 96-well plate for flow cytometry staining. iMGLs were first treated with Zombie Aqua™ dye (Fixable Viability kit, BioLegend, Cat# 423102) for 20 mins in PBS (1:1000) followed by washing with FACS buffer and centrifugation at 500g for 5 mins. iMGLs were then processed and stained with CD45-Alexa Fluor 700, CD11b-PE/Cy7 for defining iMGLs by flow cytometry. iMGLs were then run on a BD X-20 analyzer and acquired for the stained markers, and TAU-PFF ATTO 488 staining was quantified in CD45+CD11b+ iMGLs. The resulting data were analyzed with FlowJo software from BD (FlowJo 10.8.1). Bar graphs for quantitative representation of flow cytometry data were generated in GraphPad Prism 10 (version 10.0.3).

#### sTREM2 ELISA

On day 25 post-HPC generation, iMGLs were seeded into 6-well plates (500,000 cells/well) in iMGL basal medium with maintenance and maturation cytokines and fed on alternate days until reaching maturation on day 28. On day 28 post-HPC generation, supernatants from iMGL culture were harvested, protease and phosphatase inhibitors were added. Samples were analyzed for sTREM2 using a commercially available Human TREM2 ELISA Kit (Abcam, Cat#: ab224881) following manufacturer’s instructions. Briefly, samples were diluted 1:5 in sample diluent and 50 µl samples were added to each well of pre-coated microplate strips. Three biological replicates were run as technical duplicates. Plates were read using 450 nm using a BioTek Microplate spectrophotometer.

#### Proteomics

Media from iMGL cultures were analyzed by OLINK Explore 3072 platform, a high-throughput, multiplex protein immunoassay-based proximity extension assay including a protein library of 2944 protein assays ^81^. iMGL basal medium with maintenance and maturation cytokines served as negative controls for assessing background level of the proteins measured. Briefly, target proteins are bound by unique pairs of oligonucleotide-labelled antibodies. When in close proximity to one another, the oligonucleotides hybridize to form a PCR target sequence.

The resulting DNA amplicon is quantified on an Illumina NovaSeq platform using Next Generation Sequencing. Protein levels derived from cycle threshold values are directly proportional to the concentration of target protein in the sample and are reported on a log2 scale as normalized protein expression (NPX). The technical QC was passed for 2902 of the 2944 assays. Using our standard proteomics data analyses pipeline ^81^, low detection proteins were filtered out. Proteins with detectability in all of the mutant samples, all of the WT samples, or in all samples were included in the analyses (n=1490). Proteins were excluded if the average coefficient of variation (CV) was above 20% within the technical triplicates (n=1360 protein assays). We averaged the triplicate measurements for each sample and compared the expression of secretory factors between mutation carriers vs isogenic control WT iMGL cultures by paired t-tests. Significant secretory factors (p-value≤0.05) were categorized into those upregulated or downregulated and functional annotations of these factors into pathways were derived from EnrichR gene list enrichment analysis tools ^126^.

#### iMGL conditioned media treatment on iPSC-neurons

Human iPSCs from a normal, healthy control donor were engineered to introduce a dox regulatable form of Ngn2 (F12468.13.CRISPRiΔ1E11.NgN2Δ3B09) in the AAV safe harbor locus as previously described ^128^. Engineered iPSCs were characterized for pluripotency markers and stable karyotype. To differentiate iPSCs into cortical neurons, iPSCs were plated at a density of 65,000 cells per well in neural induction media (StemCell Technologies, Cat #05839) in a 96-well v-bottom plate to form neural aggregates. After 5 days, cells were transferred into culture plates. The resulting neural rosettes were isolated by enzymatic selection (Neural Rosette Selection Reagent; StemCell Technologies, Cat #05832) and cultured as neural progenitor cells (NPCs). NPCs were differentiated into neurons upon 1 µg/mL doxycycline induction (doxycycline hyclate, Sigma, cat#: D5207-10G) in BrainPhys™ Neuronal Media (STEMCELL Technologies, cat#: 05790) supplemented with BDNF (20ng/ml, PeproTech, 450-02-100µg), GDNF (20ng/ml, PeproTech, 450-10-100ug), cAMP (0.5mM, Sigma, cat#: D0627), glutamax (1X, ThermoFisher Scientific, cat#: 35050061), B-27™ Supplement (50X, 1: 100, Life Technologies, cat#: 17504-044), N-2 Supplement (100X, 1:200, ThermoFisher Scientific, cat#: 17502001), and pen/strep (ThermoFisher Scientific, 15140-122). Neurons were then replated on coverslips in 24 well plate tissue culture dishes, coated with Poly-l-ornithine (0.01%, R&D Systems, cat#: 3436-100-01; coating overnight) and laminin I (10µg/ml, R&D Systems, cat#: 3400-010-03; coating for 2 hours) at 5×10^4^ cells/well. On day 7, neuron media was removed and replaced with naïve neuronal conditioned media or conditioned media from isogenic *MAPT* WT or *MAPT* IVS10+16 iMGL cultures (400µl/well). Neurons were cultured for an additional 7 days with media changes every 2 days.

On day 14, neuronal cells were first fixed with 4% paraformaldehyde followed by three washes with 1X DPBS and permeabilized with 0.1%Triton X-100 and then incubated for 1 hr in blocking buffer (1X DPBS with 0.1% saponin, 1% BSA, and 5% FBS). Neurons were then incubated in blocking buffer containing primary antibody (see below) overnight at 4°C. After washing 3 times with 1x DPBS, secondary antibodies were added in blocking buffer for 1 hr at RT in the dark. Antibodies used for immunocytochemistry of neurons included synapsin-1(Millipore, cat #: AB1543P, 1:500), MAP2 (Abcam cat #: ab5392, 1:500), and goat Alexa Fluor conjugated secondary antibodies (Invitrogen,1:1000). Coverslips were mounted on slides with Fluoromount-G™ (Thermo Fisher Scientific, 00-4958-02). Images were acquired with an LSM980 confocal system (Zeiss) with Airyscan and a 20x or 63x oil-immersion objective lenses.

Synapse density shown in Figure 5G was quantified as the number of synapsin puncta per mm^2^ of field area (number of cells analyzed for Synapsin-1 puncta density per image field; neuron media, n=103; WT iMGL media, n=72; IVS10+16 iMGL media, n=87). Morphometric analyses were performed on 40x confocal images, 3D reconstructed in Imaris software and semi-automatically analyzed for mean dendritic length (number of neurites analyzed; neuron media, n=172; WT iMGL media, n=158; IVS10+16 iMGL media, n=105, respectively).

## Data Availability

All data produced in the present study are contained in the manuscript and/or available upon reasonable request.

## Acknowledgments

We would like to thank the research subjects and their families who generously participated in this study. We thank Torri Ball and Fabia Filipello for careful review and thoughtful discussion of the study. We thank Dr. Aimee Kao who generously provided the fibroblasts used to generate the GIH36 and GIH178 iPSC lines (supported by the Rainwater Charitable Organization). GIH36 and GIH178 were provided through the generous support of the Tau Consortium of the Rainwater Charitable Foundation. We thank Steve Lotz and members of the Neuracell Core Facility, the Neural Stem Cell Institute, Rensselaer NY for generating the GIH36 and GIH178 iPSC lines. We thank the Genome Engineering & Stem Cell Center (GESC@MGI) at Washington University in St. Louis School of Medicine for iPSC reprogramming services. This work was supported by access to equipment made possible by the Hope Center for Neurological Disorders, the Neurogenomics and Informatics Center, and the Departments of Neurology and Psychiatry at Washington University School of Medicine. Confocal images were generated on a Zeiss LSM 980 Airyscan Confocal Microscope which was purchased with support from NIMH grant MH126964 and on the Nikon AXR with NSPARC through the use of Washington University Center for Cellular Imaging (WUCCI)-Neuro supported by Washington University School of Medicine, the Children’s Discovery Institute of Washington University and St. Louis Children’s Hospital (CDI-CORE-2015-505 and CDI-CORE-2019-813), and the Foundation for Barnes-Jewish Hospital (3770 and 4642). Funding provided by the National Institutes of Health (P30 AG066444, R01 AG056293, RF1 NS110890, U54 NS123985), Hope Center for Neurological Disorders (CMK), Rainwater Charitable Organization (CMK), BrightFocus Foundation (CMK), Farrell Family Fund for Alzheimer’s Disease (CMK), and UL1TR002345. CS is supported by National Institutes of Health (NIH)/National Institute on Aging (NIA) K01AG062796. LV is supported by grant funding and consultancy/speaker fees from ZonMw (VENI grant), Amsterdam UMC (Startergrant) Stichting Dioraphte (biobank DemenTree), Olink, Lilly, Roche; all paid to her institution. Mass spectrometry work was supported by resources and effort provided by the Tracy Family Stable Isotope Labeling Quantitation Center established by the Tracy Family, R. Frimel and G. Werths, GHR Foundation, D. Payne and the Willman Family brought together by The Foundation for Barnes-Jewish Hospital. This work was also supported by cores, resources, and effort provided by Washington University Biomedical Mass Spectrometry Research Facility (NIH/National Institute of General Medical Sciences P41 GM103422). The UCSF Neurodegenerative Disease Brain Bank receives funding support from NIH grants P30AG062422, P01AG019724, U01AG057195, and U19AG063911, as well as the Rainwater Charitable Foundation, and the Bluefield Project to Cure FTD. We would like to thank Dorjan Brinja, Pascaline Akitani, and Erica Lantelme of the Flow Cytometry & Fluorescence-Activated Cell Sorting Core supported by the Department of Pathology and Immunology at Washington University in St. Louis for their assistance with flow sorting of HPCs. Diagrams were generated using BioRender.com.

## Supplemental Figure Legends

**Figure S1.**
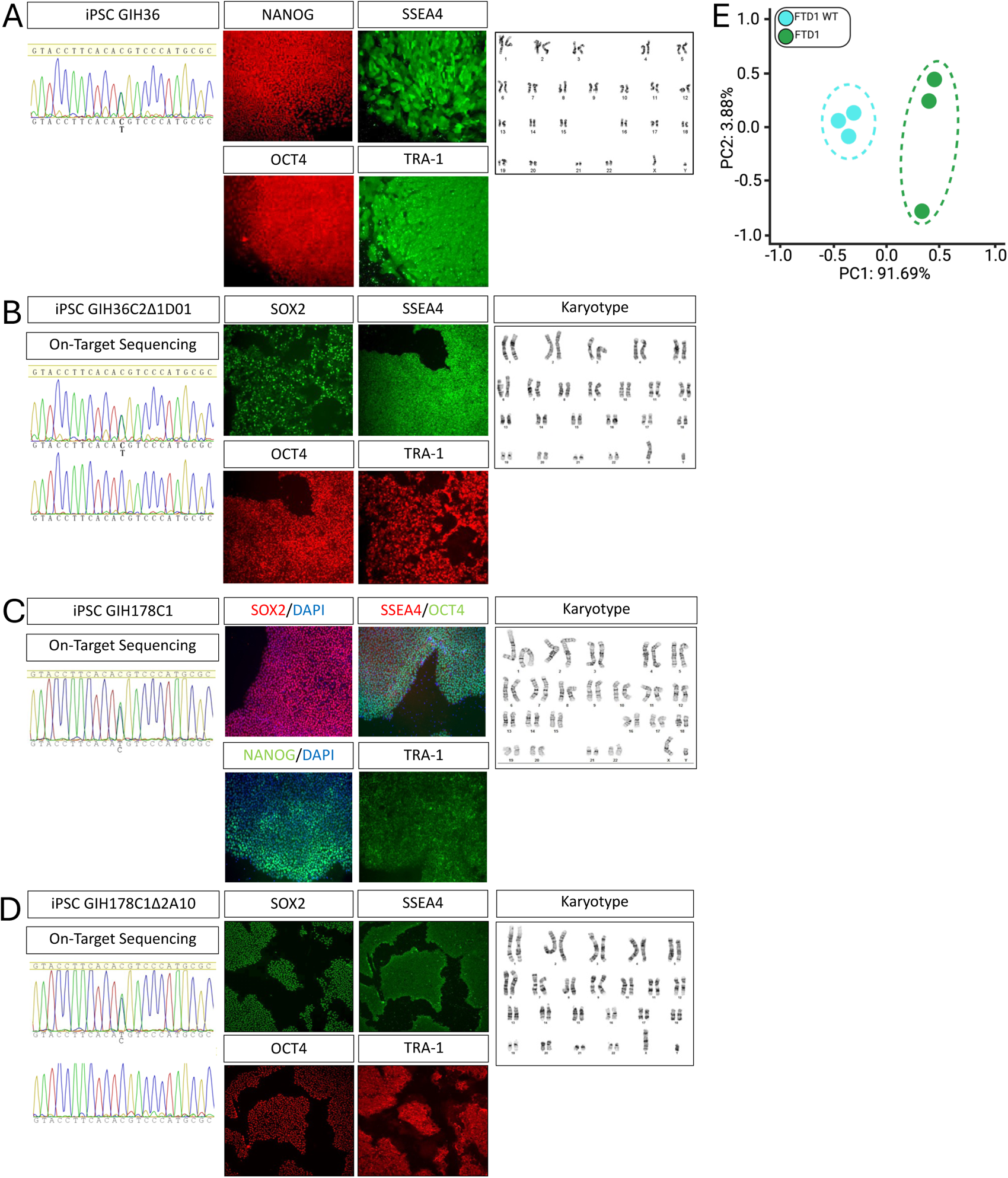
iPSC characterization and QC. A-D. iPSC characterization for *MAPT* IVS10+16 donor lines and isogenic controls. Each iPSC line used in the study was verified by Sanger sequencing. Karyotyping was performed to confirm chromosomal stability. Immunocytochemistry was performed to confirm expression of pluripotency markers. A-B. Isogenic pair, FTD1. A. GIH36, *MAPT* IVS10+16. B. GIH36C2Δ1D01, *MAPT* WT. C-D. Isogenic pair, FTD2. C. GIH178C1, *MAPT* IVS10+16. D. GIH178C1Δ2A10, *MAPT* WT. E. PCA for RNA sequencing of *MAPT* IVS10+16 and isogenic control iMGLs.

**Figure S2.**
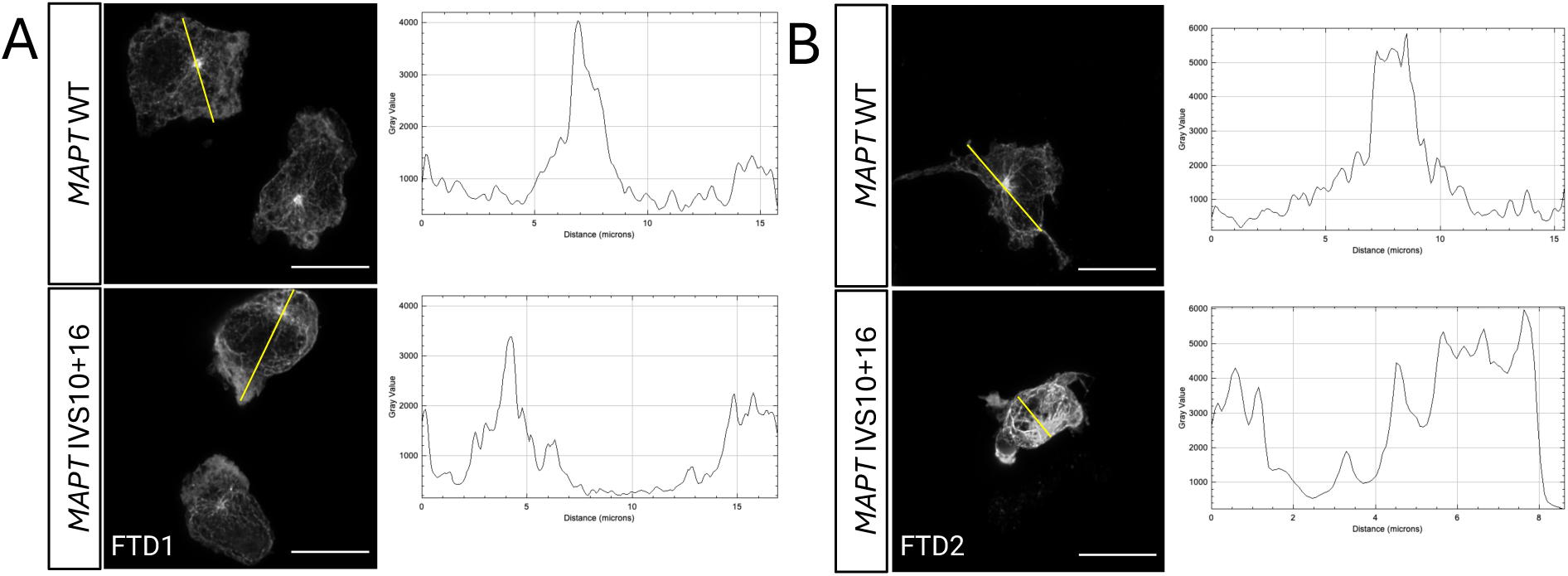
Representative images used for quantification shown in Figure 2F depicting sections drawn using straight line tool (in yellow) in ImageJ for assessing distribution of IZl-tubulin staining in FTD1 (A) and FTD2 (B) donor pair iMGLs. Alongside each donor pair images are representative histograms showing raw mean fluorescence intensity of IZl-tubulin along the length of the shown yellow line or distance along a given cell.

**Figure S3.**
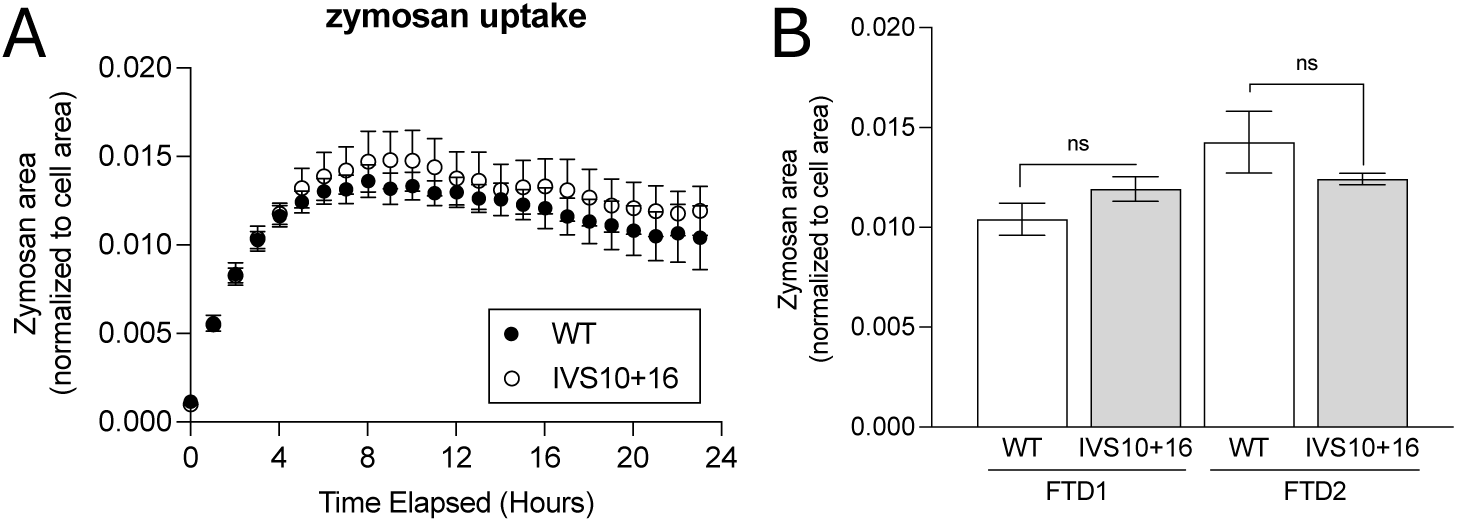
(A) Phagocytic uptake of zymosan A (S. cerevisiae) BioParticles™, Alexa Fluor™ 488 conjugate (0.5 µg/ml) in FTD1 and FTD2 donor pair iMGLs assessed by IncuCyte live cell imaging assay for a period of 24h from substrate addition. (B) 12h timepoint quantification of zymosan area normalized to microglia area are shown for FTD1 and FTD2 donor pair iMGLs. N=8 replicates/line.

**Figure S4.**
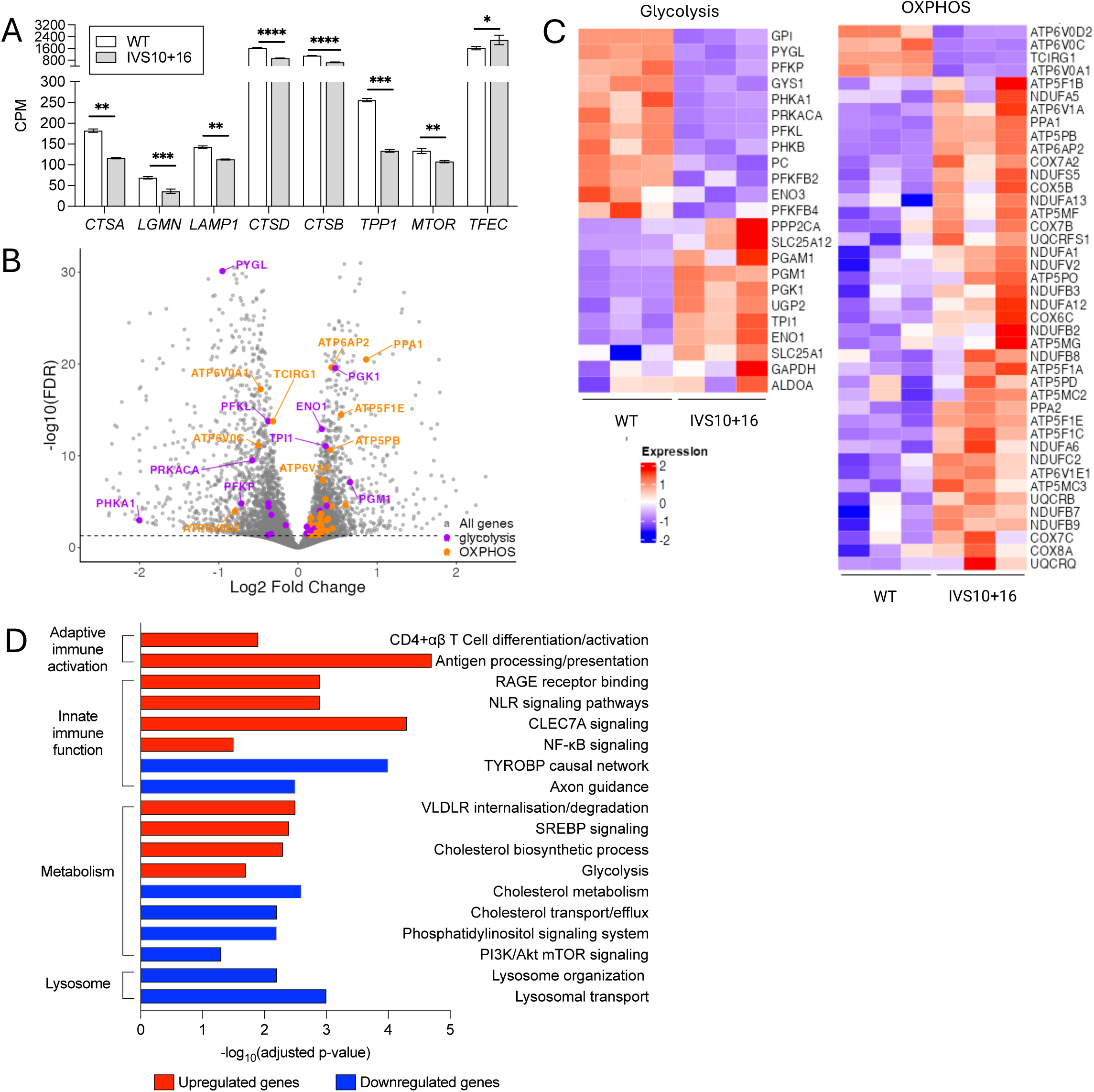
*MAPT* IVS10+16 disrupts glycolytic and oxidative phosphorylation pathways in iMGLS. A. Autophagy lysosome pathway genes are differentially expressed in *MAPT* IVS10+16 iMGLs. Normalized counts per million transcripts. *, FDR-BH <0.05; **, FDR-BH<0.01; ***, FDR-BH<0.0005, ****, FDR-BH<0.0001. B. Volcano plot revealed dysregulation of genes involved in glycolysis and oxidative phosphorylation (OXPHOS) pathways. C. Heatmap of genes involved in glycolysis and oxidative phosphorylation. D. Pathway analysis of genes differentially expressed between *MAPT* IVS10+16 and isogenic control iMGLs shows an enrichment of genes involved in adaptive immune activation, innate immune function, metabolism and lysosome function. Red bars, significantly upregulated genes. Blue bars, significantly downregulated genes. Significance defined as FDR-BH≤0.05. Three biological replicates were included for each donor.

