## Supplementary figures and images for "Cell autonomous microglia defects in a stem cell model of frontotemporal dementia"

### Supplemental Figures

#### Supplemental Figure 1

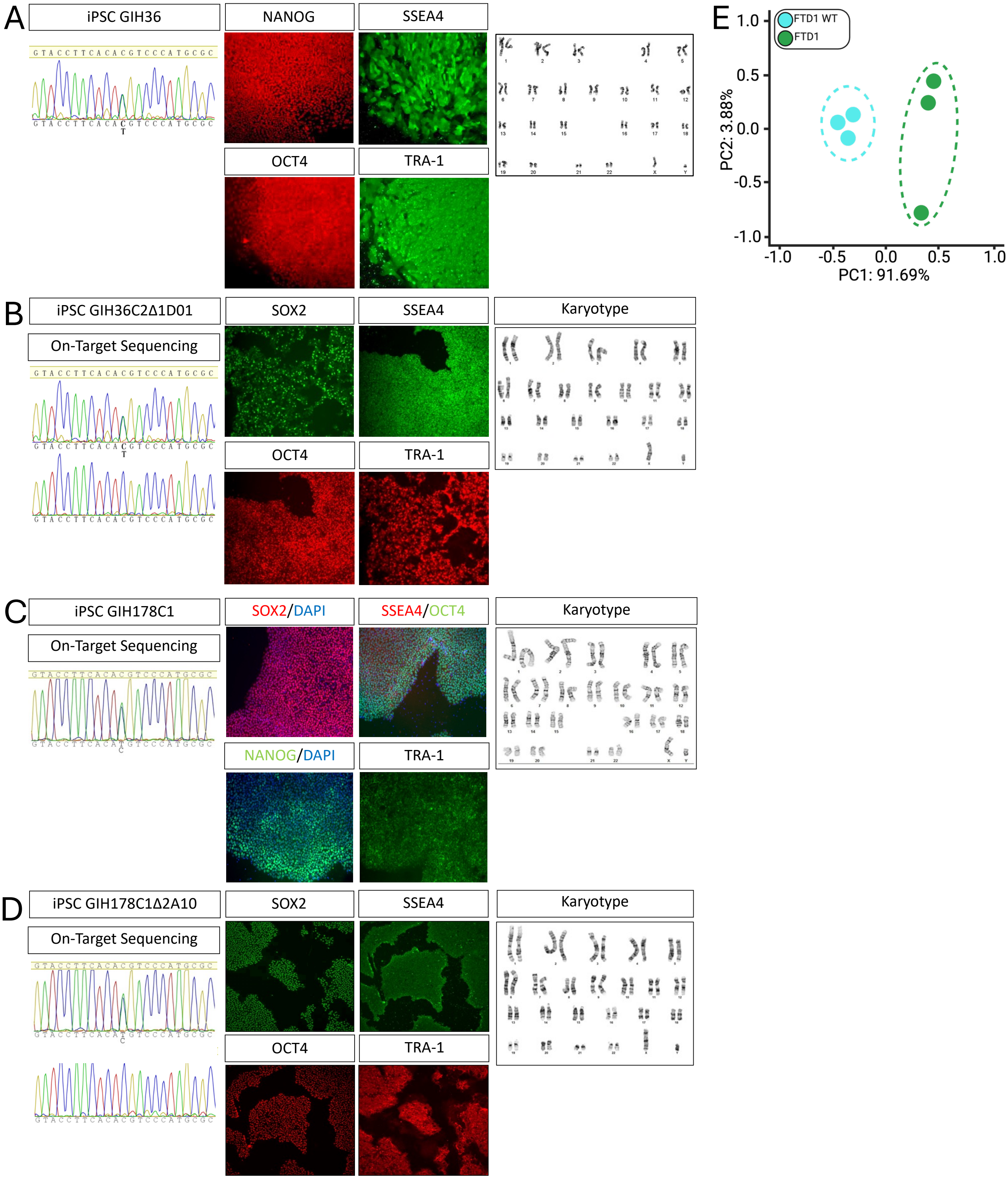

#### Supplemental Figure 2

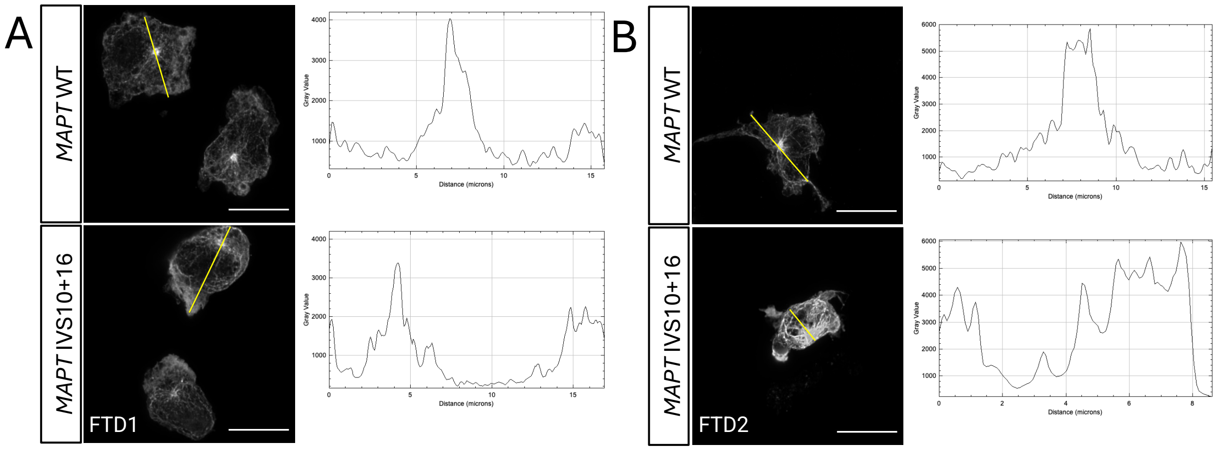

#### Supplemental Figure 3

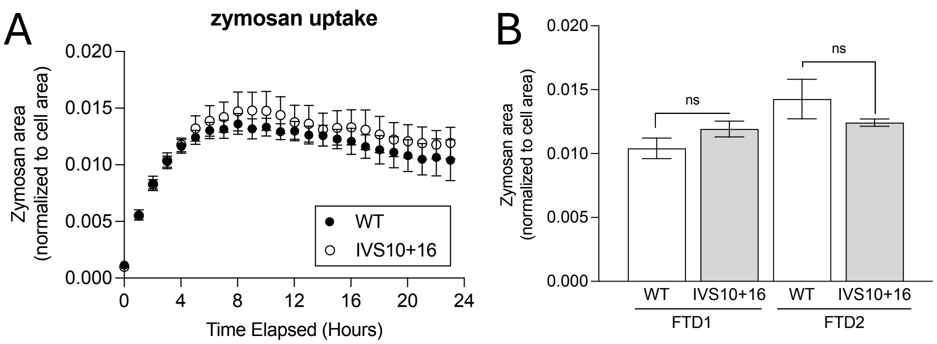

#### Supplemental Figure 4

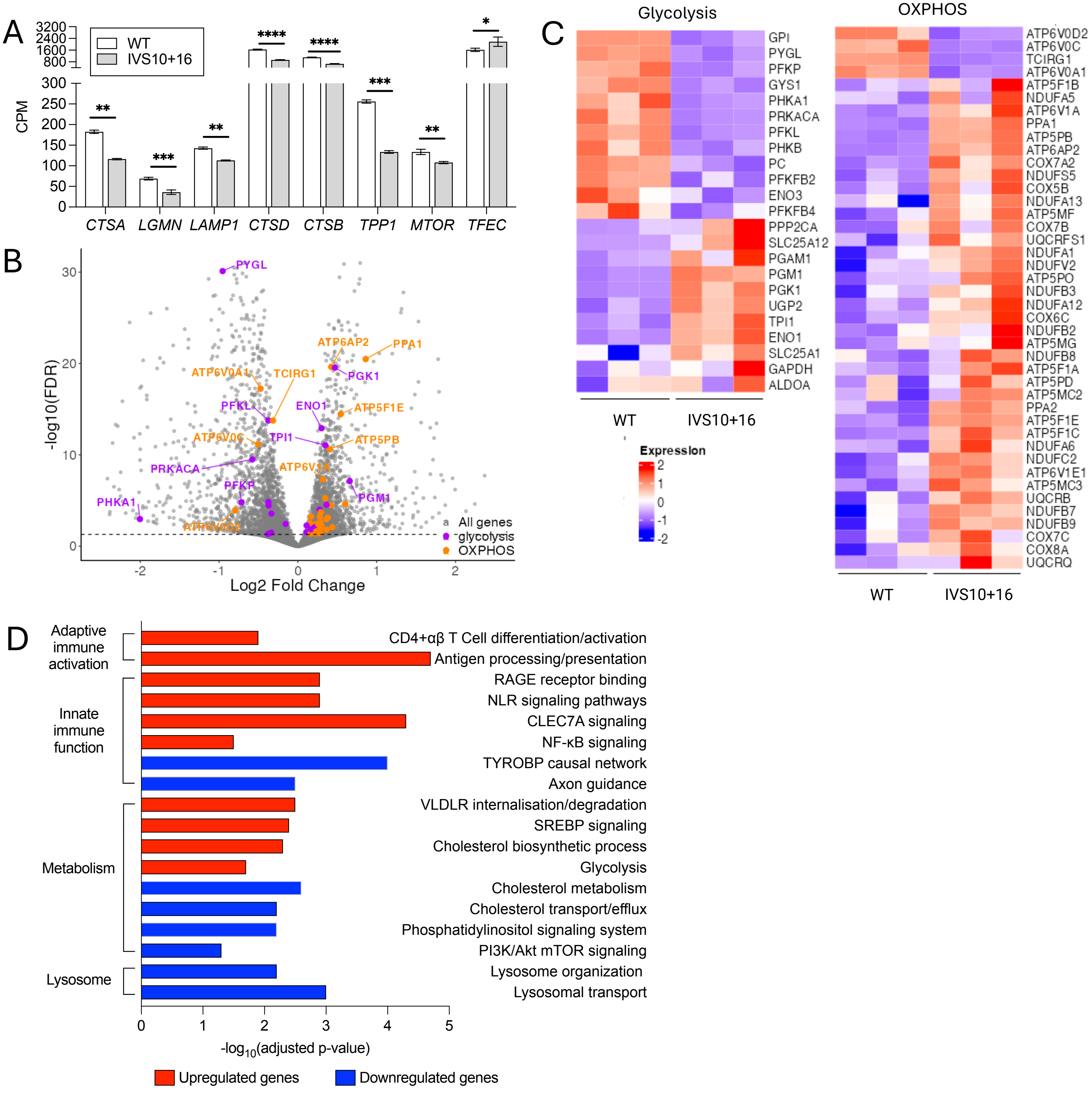
